# Immunogenic epitope panel for accurate detection of non-cross-reactive T cell response to SARS-CoV-2

**DOI:** 10.1101/2021.12.12.21267518

**Authors:** Aleksei Titov, Regina Shaykhutdinova, Olga V. Shcherbakova, Yana Serdyuk, Savely A. Sheetikov, Ksenia V. Zornikova, Alexandra V. Maleeva, Alexandra Khmelevskaya, Dmitry V. Dianov, Naina T. Shakirova, Dmitry B. Malko, Maxim Shkurnikov, Stepan Nersisyan, Alexander Tonevitsky, Ekaterina Khamaganova, Anton V. Ershov, Elena Y. Osipova, Ruslan V. Nikolaev, Dmitry E. Pershin, Viktoria A. Vedmedskia, Mikhail Maschan, Victoria Ginanova, Grigory A. Efimov

## Abstract

The ongoing COVID-19 pandemic calls for more effective diagnostic tools, and T cell response assessment can serve as an independent indicator of prior COVID-19 exposure while also contributing to a more comprehensive characterization of SARS-CoV-2 immunity. In this study, we systematically assessed the immunogenicity of 118 epitopes with immune cells collected from multiple cohorts of vaccinated, convalescent, and healthy unexposed and SARS-CoV-2 exposed donors. We identified seventy-five immunogenic epitopes, 24 of which were immunodominant. We further confirmed HLA restriction for 49 epitopes, and described association with more than one HLA allele for 14 of these. After excluding two cross-reactive epitopes that generated a response in pre-pandemic samples, we were left with a 73-epitope set that offers excellent diagnostic specificity without losing sensitivity compared to full-length antigens, which evoked a robust cross-reactive response. We subsequently incorporated this set of epitopes into an *in vitro* diagnostic ‘Corona-T-test’ which achieved a diagnostic accuracy of 95% in a clinical trial. When applied to a cohort of asymptomatic seronegative individuals with a history of prolonged SARS-CoV-2 exposure, this test revealed a lack of specific T cell response combined with strong cross-reactivity to full-length antigens, indicating that abortive infection had occurred in these individuals.

## Introduction

The COVID-19 pandemic has posed a considerable challenge for healthcare systems worldwide, necessitating the rapid development of novel diagnostic tools. RT-PCR is the gold standard assay for confirming COVID-19 infection, while serology tests are commonly used for retrospective diagnosis, assessment of vaccination efficiency, and measuring the stability of immune protection over time. Nevertheless, estimating the actual rate of infection is complicated because many infections are asymptomatic, and up to 15% of patients do not develop a humoral immune response to infection ^1–3^

T cell response can offer an independent metric of SARS-CoV-2-specific immunity in the aftermath of either COVID-19 ^4–8^ or vaccination ^9–13^. It has been demonstrated that IgG titers strongly correlate with ^14–16^ and that antibodies can provide protection even in the absence of T cells both in animal models ^17,18^ and in prospective human studies ^16,19^. Other studies have suggested that cellular immunity has a role in the context of suboptimal humoral response ^16,18,19^ or at the early stages after vaccination before seroconversion ^20,21^. It has also become clear that the humoral response gradually fades and may no longer be detectable six months post-infection ^22,23^ or -vaccination ^24^, whereas T cells persist long after exposure ^23,25,26^. Indeed, T cell responses have remained detectable for up to 17 years after infection with SARS-CoV-1 ^27^.

However, the detection of SARS-CoV-2-specific T cell response is hindered by the relatively frequent occurrence of false-positive responses in non-SARS-CoV-2-exposed individuals due to cross-reactivity to other coronaviruses ^4,7,28^. The role of this response remains controversial; some studies report that such low-affinity cross-reactive responses may contribute to a poor prognosis ^29^, while others have demonstrated that pre-existing memory T cells rapidly respond upon vaccination ^30^ and that the expansion of cross-reactive T cells is associated with mild disease ^31^ and may explain asymptomatic infections ^32^. Nevertheless, it is important to distinguish between cross-reactive and COVID-19-specific T cell responses. To date, several kits for *in vitro* detection of T cell response have been proposed based on ELISpot/Fluorospot technology ^33–36^, high-throughput sequencing (HTS)-based detection of T cell receptor (TCR) sequences ^37^ and measuring cytokine production in whole blood ^38^. Most of these exploit custom peptide sets that were bioinformatically selected to minimize cross-reactivity, but to the best of our knowledge, these were not experimentally validated on pre-pandemic samples.

Previous studies have predicted ^39^ and experimentally confirmed ^37,40–42^ numerous SARS-CoV-2 T cell epitopes. Some researchers have focused on the properties of individual epitopes, such as the diversity and repertoires of specific T cell receptors and structural aspects of epitope recognition ^7,43–48^. Others have aimed at characterizing the response to sets of epitopes ^26,48–50^. Nevertheless, the employment of different assays hinders direct comparison, and the limited cohort size and number of epitopes tested per study have left essential questions pertaining to the immunodominance of individual epitopes—and for some epitopes, their HLA restriction—unresolved.

Understanding patterns of immunodominance of SARS-CoV-2 epitopes could guide future vaccine development. For example, ORF1ab- and ORF3a-derived epitopes seem to be more immunogenic than other components of the viral proteome—including the S glycoprotein—in individuals bearing HLA-A*01:01, which is common in the European population ^37,40,49^. Moreover, although several studies have demonstrated that the total magnitude of CD8^+^ and CD4^+^ T cell response in people vaccinated with existing S protein-based vaccines is on par with or even surpasses that of patients who have recovered from infection ^9–11,13^, it remains unclear whether the spectrum of recognized epitopes is the same in both groups. Given that increased diversity of recognized epitopes is known to correlate with better outcomes in some other viral infections, such as with hepatitis B virus ^51^, it is important to profile T cell immunity—including the landscape of recognized epitopes—in vaccinated individuals and patients after natural infection with SARS-CoV-2.

In the present study, we aimed to systematically characterize a pre-selected set of 118 SARS-CoV-2 epitopes presented by common HLA-I and -II alleles. In sharp contrast to full-length antigens, the selected epitopes did not induce a response in pre-pandemic samples, with the exception of two HLA-II-restricted peptides. We confirmed immunogenicity for 75 epitopes, and HLA restriction for 49 of them, including nine HLA-I epitopes that had previously displayed ambiguous binding. We further demonstrated that seven epitopes are presented by more than one HLA allele. 26 epitopes were immunodominant, meaning they were identified in at least 50% of patients with the restricting HLA allele. Based on these findings, we designed the ELISpot-based *in vitro* diagnostic ‘Corona-T-test’, which is designed for specific detection of COVID-19- or vaccine-induced—but not cross-reactive—T cell response to SARS-CoV-2. This test demonstrated 95% accuracy in a clinical trial of 69 vaccinated individuals, 50 COVID-19 convalescent patients (CPs), and 101 unexposed donors. We subsequently used this test to study a cohort of asymptomatic seronegative individuals with a history of prolonged SARS-CoV-2 exposure, and observed a lack of specific T cell response and substantial cross-reactivity to full-length antigens, indicating abortive infection in this cohort. Finally, we demonstrated that individuals vaccinated with the two-component Gam-COVID-Vac adenoviral vaccine (Sputnik V) maintained a strong, broad, and diverse CD8^+^ T cell response at a median of 66 days after the first injection, with a significantly higher number of recognized S-derived epitopes but negligible CD4^+^ reactivity in comparison to CPs.

## Results

### Identifying a set of putative immunogenic and non-cross-reactive T cell epitopes

To assemble a set of peptides, we collected available information on SARS-CoV-2 T cell epitopes ^7,37,40–42,48,50^, as depicted in **Fig.1A**. The selected peptides were derived from both structural and non-structural SARS-CoV-2 proteins, based on their high immunogenicity in CPs and low immunogenicity in non-exposed individuals as well as their predicted binding to one or several common HLA alleles across the European population. The final set included 94 putative HLA-I binders (*i.e.,* MHC-I peptides) and 24 putative HLA-II binders (*i.e.,* MHC-II peptides), where each peptide was predicted to bind on average to four HLA-I and five HLA-II alleles, respectively (**Table S1**, **Fig. 1A**). We did not observe a difference in the homology scores of the selected peptides and a set of other SARS-CoV-2 immunogenic epitopes annotated in the Immune Epitope Database (IEDB) relative to common cold coronaviruses (**Fig. S1A-D**). To estimate the theoretical coverage of the population for this set of peptides, we evaluated the frequency distribution of the restricting HLA alleles among HLA-typed individuals in the local bone marrow donor registry (n = 2,210). Only a single person (0.05%) had none of the alleles that were predicted to present these MHC-I or MHC-II peptides (**Fig. 1B**), indicating the designed peptide set offers sufficient predictive sensitivity.

**Figure 1.**
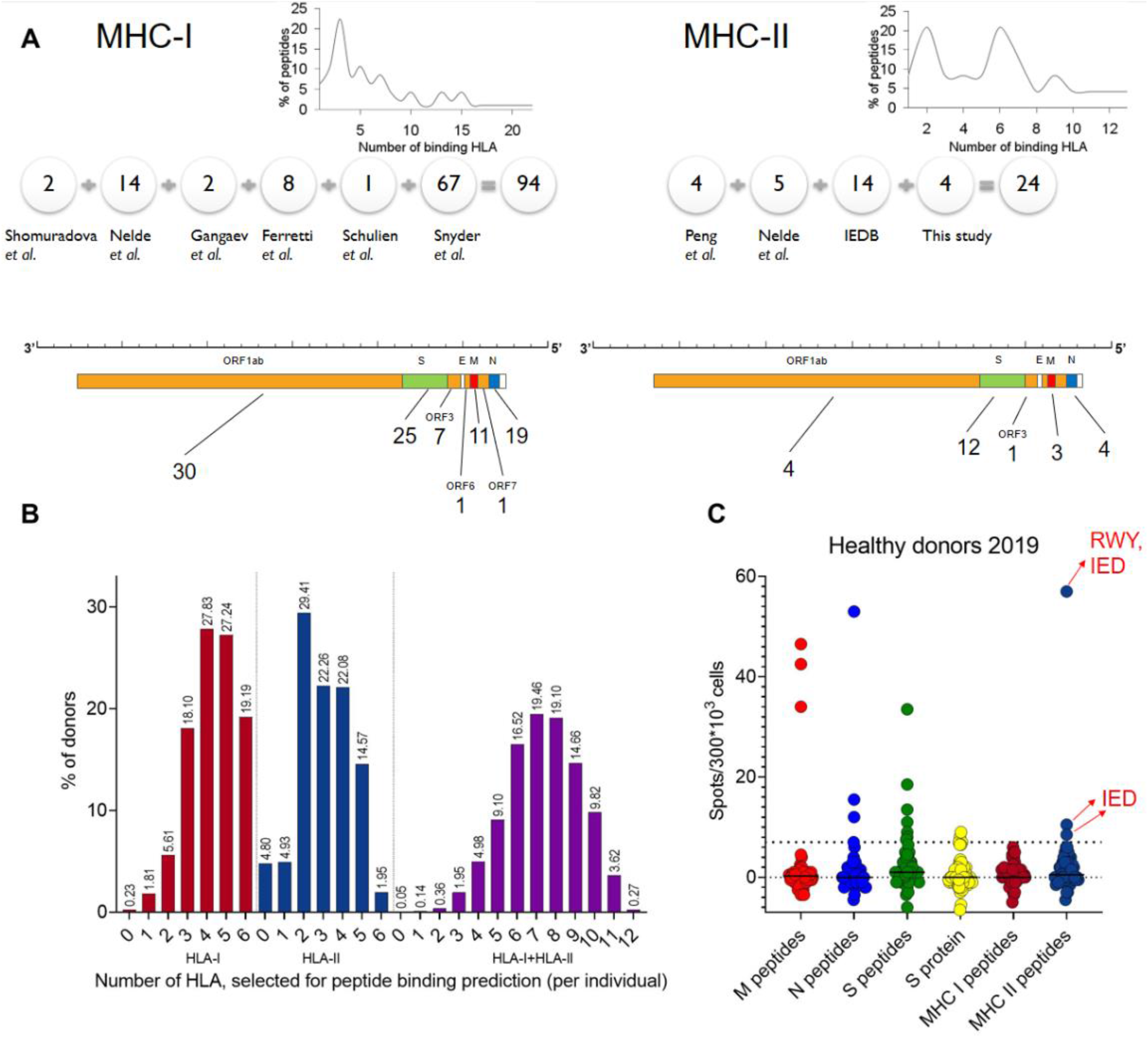
Characteristics of the peptide set and strong cross-reactive responses to full-length antigens in unexposed healthy donors. (A) Number of epitopes selected from each indicated publication (detailed in **Table S1**) for the MHC-I (left) and -II (right) sets. The distribution of the peptides according to the number of HLA that they bind is shown at top. The x-axis displays the number of predicted binding alleles per peptide. The y-axis shows the percentage of peptides that bind to a given number of alleles. Numbers below the SARS-CoV-2 genome schematic indicate the number of peptides derived from each gene. (B) The number of HLA class I (left) and II (middle) alleles alone or in combination (right) that are predicted to bind at least one peptide from the set per individual among 2,210 donors from the bone marrow registry. (C) Antigen response among our healthy donors 2019 (HD2019) cohort (n = 52). Two cross-reactive peptides from the MHC-II peptides are marked with red arrows. Dots represent the mean of two duplicates with negative control subtracted. The positive threshold (7 spots) is indicated by the dotted line.

### Full-length antigens induce a cross-reactive response compared to the selected peptides

We compared the specificity for pools of peptides spanning the full length of the various SARS-CoV-2 structural proteins—S, nucleoprotein (N), and membrane protein (M)— with that for the MHC-I and -II peptide sets using a cohort of pre-pandemic healthy donor samples (HD-2019; n = 52) and measuring the interferon ɣ (IFNɣ) response by ELISpot. Ten donors produced a positive (≥7 spots) T cell response to any of the peptide pools (S, N, or M) (**Fig.1C**) or to recombinant S protein. Three of these donors also had a positive response to the set of MHC-II peptides (**Fig. S1E**). Using matrix pools (see Methods), we identified two cross-reactive MHC-II peptides (RWY from N protein and IED from S protein; **Fig. 1C**, **S1F**; sequences are given in **Table S1**). The high frequency of cross-reactive responses induced by the S, N, and M peptide pools or by recombinant S protein makes these targets ill-suited for measuring SARS-CoV-2-specific T cell response.

### Diverse response in convalescents and MHC-I-focused response in vaccinated

We next analyzed the response to S, N, and M peptides and MHC-I and MHC-II peptides in cohorts of COVID-19 convalescent patients (n = 51, CP) and Sputnik V-vaccinated individuals (n = 45, Vac). Full information on these cohorts is provided in **Table S2**. We excluded the two cross-reactive peptides identified above (RWY and IED) to create a new ‘MHC-II cross^—^ peptides’ set; the initial MHC-II set will subsequently be referred to as ‘MHC-II cross^+^’. In agreement with recently published data ^13,35^, we observed that Vac individuals demonstrated a greater response to S peptides than CP, while the response to N and M peptides in Vac was non-existent. Both cohorts demonstrated comparable responses to the MHC-I set (**Fig. 2A**), although peptides derived from S protein accounted for only 27% of that set.

**Figure 2.**
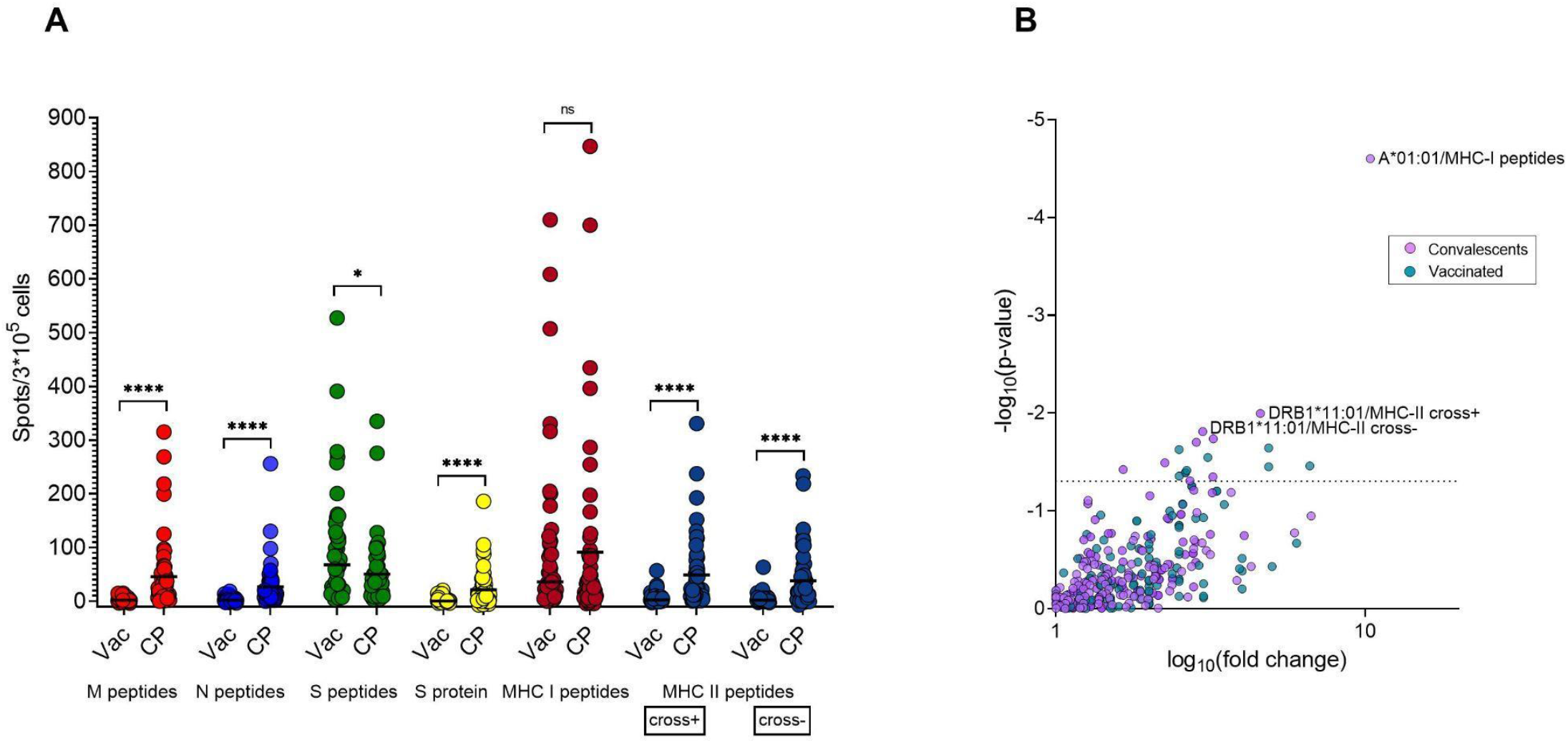
Response to MHC-II peptides differs significantly in Vac and CP donors. (A) Response to the indicated antigens as measured by ELISpot for Vac (n = 43) and CP (n = 51). Each dot shows mean of two wells after subtracting negative control. Mann-Whitney test (S peptides – p = 0.013; MHC-II peptides, recombinant S protein, N and M peptides – p < 0.0001). (B) Volcano plot shows the effect of a particular HLA allele on response to the same peptide sets and antigens. The x-axis denotes the decimal logarithm of the ratio of the median response among HLA carriers to that of individuals without the HLA. The y-axis denotes the negative decimal logarithm of the p-value. The three most significant associations are annotated. P-value = 0.05 is depicted by the dotted line (Fisher exact test).

Surprisingly, individuals in the Vac group demonstrated a significantly weaker response to MHC-II peptides in comparison to CP (**Fig. 2A**). We hypothesized that the MHC-II peptide set, although it included multiple S-derived peptides, was skewed toward immunogenic peptides from the other antigens, N and M. We tested this by selecting the eight CP donors with the strongest response to the MHC-II peptides (>50 spots) and calculating the ratio of the number of spots in wells containing peptides from the MHC-II cross^—^ set to the number in wells containing MHC-II peptides derived from S protein (excluding IED peptide). We observed a detectable response to the latter set in just one donor, accounting for ∼8% of the response to MHC-II cross^—^ peptides (**Fig S2A**). This suggests that S-derived MHC-II epitopes might be non-immunogenic or evoke a low-frequency T cell response that is barely detectable without *ex vivo* expansion. We also observed negligible response in the Vac cohort to the recombinant S protein (**Fig. 2A**) in comparison to CP, where this response was more strongly correlated with the response to MHC-II peptides than to MHC-I peptides (**Fig. S2B**). This probably reflects the predominant presentation of the recombinant protein by MHC-II pathway. We tested whether time influenced the antigen response within our sampling period, and did not observe significant association with the recombinant S protein, MHC-I, or MHC-II peptides (**Fig. S2C–H**).

Based on the sum of positive spots in wells with MHC-I and MHC-II cross^—^ peptides (‘MHC-I + II’) we made the surprising observation that the CP cohort demonstrated comparable responses to our limited set of peptides and to the S, N, and M peptides (**Fig. S3A**). This effect was replicated in the Vac cohort, where the response to the MHC-I + II set was not different from the response to the S peptides. Notably, several Vac donors demonstrated even greater responses to the MHC-I + II set in comparison to S peptides. We next assessed the contribution of cross-reactive peptides IED and RWY to the total response (**Fig. S3B**). We observed a significant impact of these peptides (> 30 spots) in three CP donors, but this response was only present in patients with high reactivity to other MHC-II peptides. Patients with low responses to these cross-reactive peptides demonstrated virtually no difference between MHC-II cross^+^ and cross^—^ sets, and we therefore do not expect lower sensitivity due to the exclusion of the IED and RWY peptides.

We next examined the effect of the presence of a particular HLA on the magnitude of response to different antigens (**Fig. 2B**). For CP donors, we observed a very strong association between the presence of HLA-A*01:01 and the number of spots in response to MHC-I peptides. Indeed, most of the HLA-A*01:01-restricted peptides, which we confirmed as immunodominant, were derived from viral ORFs and thus could not evoke a response in the Vac cohort. This suggests that HLA-A*01:01 may be associated with increased response to ORF-derived epitopes (**Fig. S3C**). For the other HLA alleles, we observed significant variability in the responses. As an example, CP donors p2037 and p2034, who had detectable responses to MHC-I peptides (17.5 and 30 spots), exclusively carried A*02:01 out of all the HLA-I alleles that were confirmed to present immunogenic peptides (*i.e.,* ‘confirmed HLA’). In contrast, three other CPs bearing A*02:01 alongside other confirmed HLA-I alleles demonstrated only a negligible response (0; 5; and 7 spots). The increased response to MHC-II peptides that we observed in carriers of DRB1*11:01 may be associated with higher immunogenicity of some peptides in the context of this particular allele. We also compared differences in the prevalence of common HLA alleles (>10% phenotypic prevalence among bone marrow donors) between groups and observed that the CP cohort included fewer patients with B*07:02/C*07:02 (**Fig S3D**) in comparison to either bone marrow donors or the Vac cohort.

### The selected set of peptides demonstrated high diagnostic accuracy

We then assessed the diagnostic accuracy of the selected peptide set. For Vac donors, surprisingly, MHC-I + II peptides demonstrated a very similar area under the curve (AUC) compared to S peptides (0.96 vs. 0.98), suggesting that the limited number of S protein-derived peptides is sufficient for detecting T cell response here (**Fig. 3A**). In contrast, the MHC-I + II peptides discriminated CP from HD-2019 better than any of the peptide pools covering full-length antigens (**Fig. 3B**), providing the same sensitivity (94%) with better specificity (S: 88%, cutoff 8 spots vs. MHC-I + II: 94%, cutoff 7.5 spots) and a higher AUC (0.99 vs. 0.97). In the CP cohort, the lack of specificity was more prominent when we analyzed the sum of spots in wells with M, N, and S peptides (**Fig. 3C**); this reduced specificity could not be fully mitigated by application of the logistic regression model based on the spot values for the S, N, and M peptides. However, the wide confidence intervals of AUC values do not allow us to assess statistical significance for these differences.

**Figure 3.**
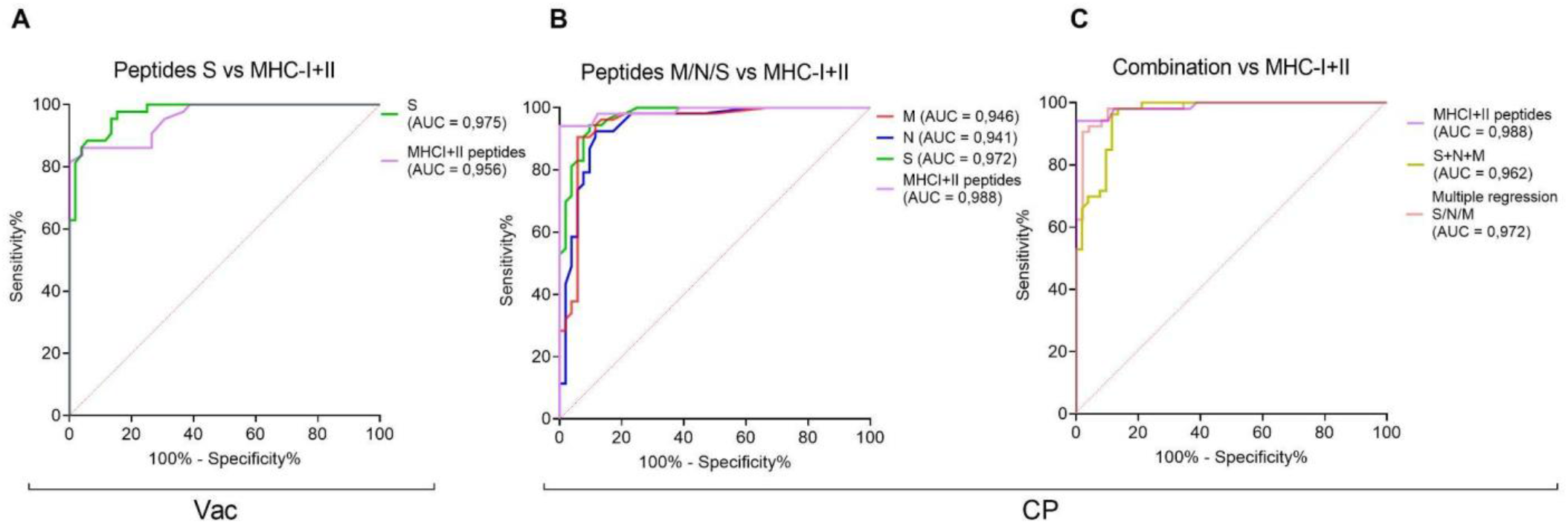
MHC-I + II peptides demonstrate greater diagnostic accuracy in CPs in comparison to the S, N, M peptides. (A) Receiver operating characteristic (ROC) curves for MHC-I + II peptides vs. S peptides in Vac vs HD-2019 samples. (B) ROC curves for MHC-I + II peptides vs. S, N, or M peptides in CP vs HD-2019 samples. (C) ROC curves for MHC-I + II peptides vs. the sum of S, N, and M peptides and vs. multiple regression model, incorporating S, N, and M peptides in CP vs HD-2019 samples. Three HD-2019 donors with cross-reactive responses were excluded from the ROC analysis for MHC-I + II peptides.

### Profiling HLA presentation of MHC-I and -II epitopes

In order to systematically analyze the immunogenicity and HLA restriction of each epitope in our set, we performed a short-term memory T cell expansion assay. Peripheral blood mononuclear cells (PBMCs) from each donor were stimulated with the complete set of MHC-I and -II peptides, split to five ‘Expansion pools’. To reduce the number of individual peptides tested in each assay, we assessed the response to smaller pools of ∼5 peptides each using IFNɣ ELISA. When a response to one of these ‘ELISA pools’ was detected, the expansion was individually tested for reactivity to the individual peptides in that pool, although only peptides that bound to that individual’s HLA alleles were tested. **Table S3** summarizes all of the epitopes that were tested as part of ELISA pools or individually. We observed at least one response to 59 of 94 MHC-I epitopes, and at least two responses to 47 epitopes. The latter epitopes were selected for HLA association analysis. Using the Fisher exact test, we identified the association with a particular HLA for 36 of the 47 immunogenic MHC-I epitopes (**Fig. 4A**). Of those, 22 epitopes were exclusively immunogenic in carriers of the associated HLA allele, with no “off-HLA” response. For four peptides, one off-HLA response was detected, and ten peptides demonstrated more than one off-HLA response. For this latter set of peptides, we searched for a second allele association that covered most of the off-HLA responses (**Fig. S4A**). For the remaining 11 peptides that did not demonstrate association with a particular HLA allele, we sought associations with a combination of two HLA alleles. We identified such a combination for two peptides (ADA and KAY, **Fig S4A**). We observed no significant association for the remaining nine peptides, suggesting that they may bind three or more alleles (*e.g.,* NRY presumably binds B*08:01, C*04:01, and C*06:02; **Table S3**) or are insufficiently immunogenic. The SSP peptide could not be unambiguously assigned, as it demonstrated the association with either C*04:01 (**Fig. 4A**) or B*35:02 and B*35:01 (**Fig S4A**). We also detected two MHC-I epitopes within predicted MHC-II epitopes.

**Figure 4.**
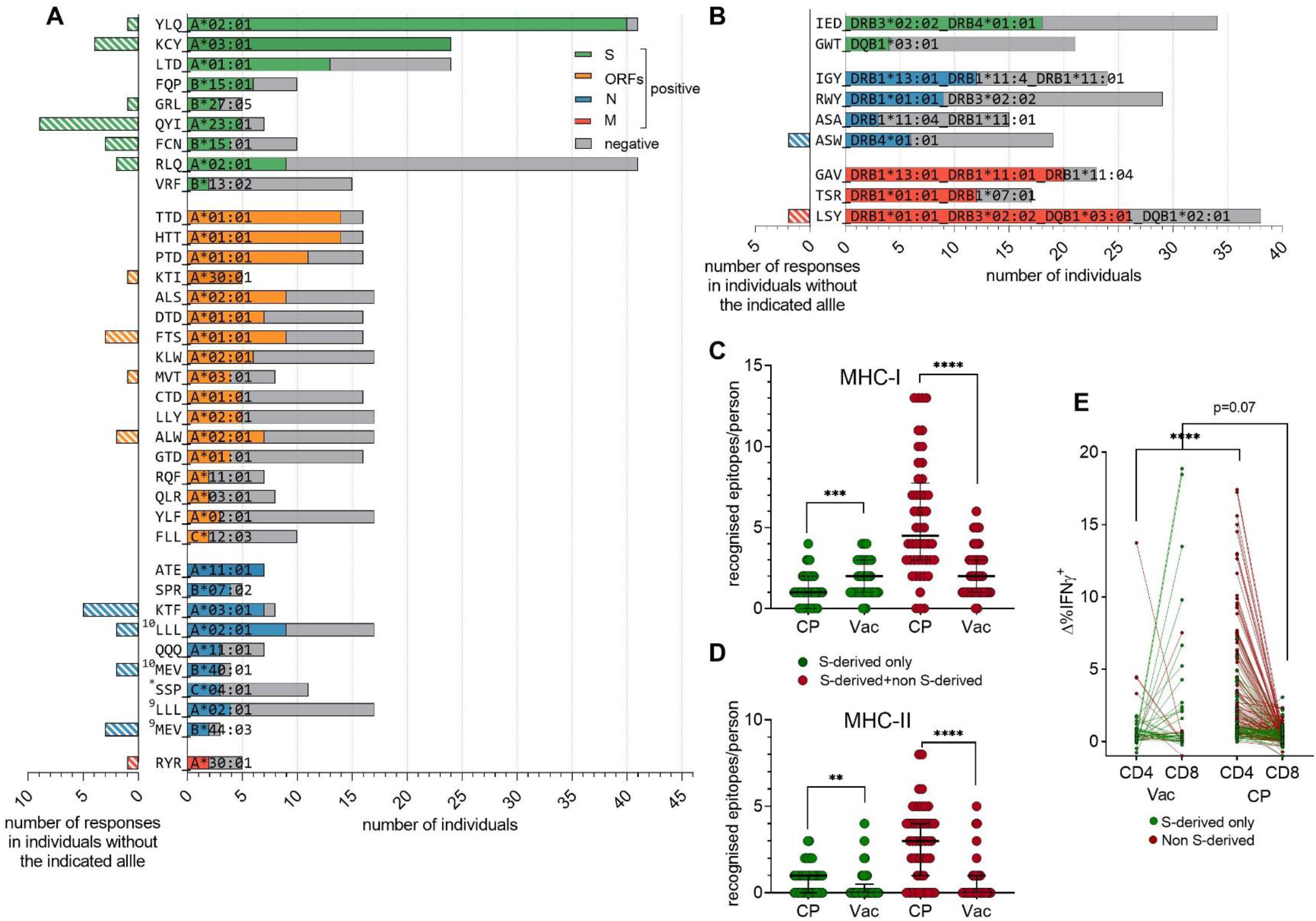
T cell response to the particular MHC-I and MHC-II epitopes in CP and Vac. Immunogenicity in carriers of various HLA-I (A) or HLA-II (B) alleles. For the righthand side of plots, grey bars indicate the number of tested carriers, colored bars indicate the number of responses. Colors indicate source protein, with three-letter amino acid codes at left. Superscript numbers indicate peptide length, and asterisk denotes a peptide with an ambiguous HLA association. Lefthand side of plots shows the number of responses in donors without the indicated HLA. For HLA-I, the association between the response and a single HLA allele is shown; for HLA-II, the best associations (including associations with several HLAs) are shown. Fisher exact test, p-values < 0.05 were considered significant (exact p-values are specified in **Table S4**). (C, D) Number of MHC-I (C) and MHC-II (D) epitopes from S or any viral protein recognized per individual for the CP and Vac cohorts. (E) Flow cytometric analysis of the phenotype of T cells responding to MHC-II peptides. Plot shows the difference in the % of CD4^+^ or CD8^+^ IFNɣ^+^ T cells between peptide-stimulated cells and negative controls.

We observed at least one response for 16 of 24 MHC-II epitopes. For the 13 epitopes demonstrating at least two CD4^+^ responses, we could not detect any apparent association with a single HLA-II allele (**Fig. S4B**). After searching for associations with a combination of two HLA alleles, several peptides still demonstrated multiple off-HLA responses (**Fig. S4B**), suggesting an association with three or even four HLA alleles. For these peptides, we present the final HLA association assignment that explained most of the responses (**Fig. 4B**). Statistics and association patterns are presented in **Table S3**. HLA alleles with a validated association with at least one immunogenic peptide are referred to as “confirmed HLA.”

Moreover, 19 MHC-I and five MHC-II epitopes were immunodominant, in that they were recognized in at least 50% of patients with the restricting HLA alleles. The most dominant MHC-I epitopes from S protein were YLQ/A*02:01 and KCY/A*03:01, with nearly 100% response; in contrast, KCY produced a response in only four out of 12 A*11:01 carriers. We also noted that QYI generated higher immunogenicity with A*23:01 in comparison to the already observed association with A*24:01 (Rowntree, 2021). Among ORF-derived epitopes, TTD/A*01:01, HTT/A*01:01, and KTI/A*30:01 elicited the most frequent response. ATE/A*11:01, SPR/B*07:02, and KTF/A*03:01/A*30:01 from N protein also showed a dominant response. In contrast to KCY, KTF was equally immunogenic both in A*03:01 and A*30:01. Assessment of immunodominance for MHC-II epitopes was hindered by their promiscuous binding, but IED, GAV, TSR, LSY, and IGY may be considered immunodominant in the context of all of the HLAs they bound.

### Immune response in vaccinated was skewed towards MHC-I epitopes

Unsurprisingly, expanded T cells from the CP cohort recognized a higher median number of MHC-I epitopes (four) than those from Vac donors (two), where the response was limited to S protein (**Fig. 4C**). However, this decrease in the number of recognized epitopes was not accompanied by a decrease in overall response magnitude (see **Fig. 2A**), suggesting a more robust response per epitope. At the same time, the number of recognized MHC-I epitopes from S protein was significantly higher in the Vac cohort (two) than in CP (one) (**Fig. 4C**). In contrast, the Vac cohort exhibited significantly fewer recognized MHC-II epitopes per person, both for all MHC-II epitopes (median 3 vs. 0) or S-derived epitopes (median 1 vs. 0; **Fig. 4D**). Using flow cytometry, we confirmed that most of the responses to S-derived epitopes were mediated by CD8^+^ rather than CD4^+^ cells (**Fig. 4E**, **Fig. S3C**). Indeed, the profile of recognized MHC-II epitopes was clearly different in Vac and skewed towards the recognition of LQT and QQL peptides, predominantly by CD8^+^ T cells (**Fig. S3C**). We observed a strong prevalence of either B*40:01 or B*44:03 among the CD8^+^ responders to QQL (four out of five). AEIRASANL (a 9-mer derived from the 15-mer QQL) was predicted to be a strong binder for both alleles, which may explain CD8^+^ reactivity to this peptide. All responders to this peptide belonged to the Vac cohort, although the frequency of B*44:03/B*40:01 was similar in the CP group (**Table S2**). Among eight responders (including six Vac) to the LQT epitope, we observed seven carriers of either A*23:01 or A*24:02. We believe these responses are due to the LQT-derived 9-mer TYVTQQLI, which is predicted as a strong binder for both alleles but seems to be more immunogenic in A*23:01 (tree our of seven responses) than in A*24:02 (four out of 23 responses). One more MHC-I epitope evoked response in two Vac individuals, but in none of the CP individuals (VRF/B*13:02) despite the similar number of B13:02 carriers. These data suggest that the observed higher number of recognized MHC-I S protein-derived epitopes per person in Vac compared to CP is either a consequence of a wider repertoire of recognized MHC-I S-derived epitopes or better detection with our readout due to a higher frequency of specific T cells.

In contrast, although T cells specific to S protein-derived MHC-II peptides could not be detected with ELISpot among PBMCs from either Vac or CP individuals (see **Fig. 2A, S2A**), they were readily detected after *ex vivo* expansion in CP but not in Vac samples (**Fig. 4D**). We did not observe any difference in the prevalence of frequent HLA-II alleles between CP and Vac groups (**Fig. S3D**), explaining the striking difference in CD4^+^ T cell response. We hypothesize that the frequency of S protein-specific CD4^+^ T cells in the Vac cohort is lower in comparison to CP. And in contrast to MHC-I epitopes, the absence of the other immunogens besides S protein does not widen the CD4^+^ response to the S protein, at least within the given timeframe of sampling. We also observed responses to non-S-derived epitopes in 10 Vac donors. In two, we observed response exclusively to cross-reactive RWY or SPR epitopes, while eight responded to two or more non-S protein epitopes, suggesting prior infection.

### Clinical confirmation of the diagnostic accuracy of the final peptide set

We designed and manufactured an ELISpot-based *in vitro* diagnostic ‘Corona-T-test’ for detection of SARS-CoV-2-specific T cells, which included the above-identified combination of peptides (designated here as “MHC-I + II_IVD”; see Methods). We performed a clinical trial in which we enrolled three independent cohorts of vaccinated (Vac_trial, n = 69), convalescent (CP_trial, n = 50), and healthy but unvaccinated individuals (HD-2021, n = 101). We measured the T cell response in parallel with the Corona-T-test and by stimulation with MHCI+II_IVD peptides followed by intracellular IFNɣ staining. As expected, intracellular INFɣ produced a low signal-to-noise ratio, and multiple patients with a robust ELISpot response were not positive based on intracellular IFNɣ staining (**Fig. S5A**). Using flow cytometry, we confirmed that the CD8^+^ response to MHC-I + II_IVD was higher than the CD4^+^ response in the Vac_trial and CP_trial cohorts (**Fig. S5B**), which is consistent with our previous ELISpot data (**Fig. S3A**). We did not observe a correlation between sampling time and humoral or T cell response in the Vac_trial cohort (**Fig. S5C–E**), which is probably due to the short timeframe that had elapsed since the boost vaccination (7-21 days). For CP individuals, we observed a weak association between sampling time and T cell or humoral response (**Fig. S5E–F**). Consistent with the previous results, our Corona-T-test demonstrated high overall accuracy in discriminating HD-2021 from Vac_trial (AUC = 0.98) and CP_trial (AUC = 0.98) (**Fig. 5A, B**) individuals, with 96.4% sensitivity, 93.5% specificity, and 95% accuracy. Several false positives in the HD-2021 might be explained either by the enrolment of asymptomatic convalescents without detectable antibody response or by the presence of cross-reactive epitopes in our MHCI+II_IVD peptide set. HLA-genotyping of the responders in the HD-2021 cohort did not reveal any obvious HLA bias (*e.g.*, high B*07:02 prevalence), most likely excluding the possibility that a single cross-reactive epitope caused the occasional false-positive responses.

**Figure 5.**
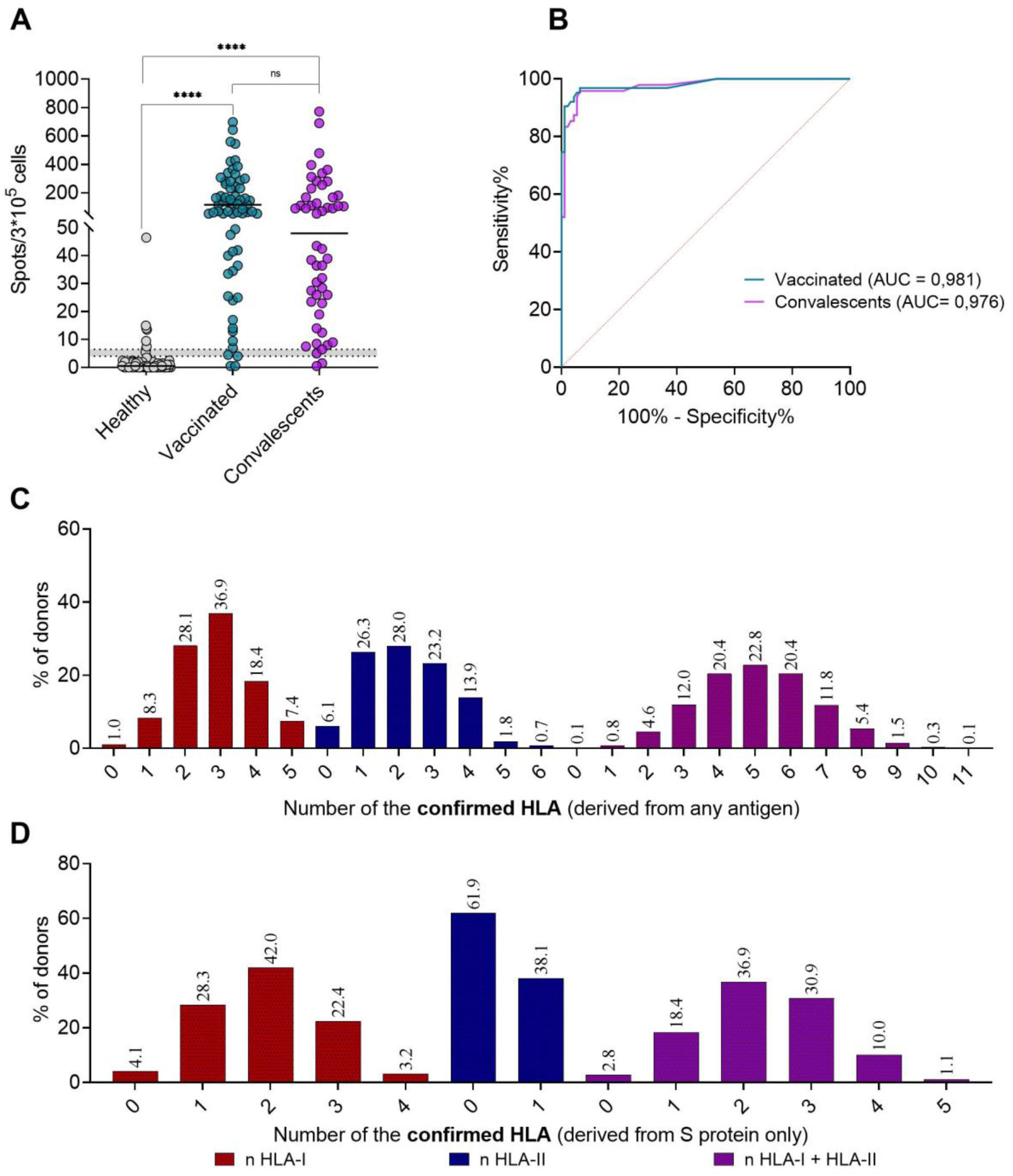
Clinical trial confirmed the high accuracy of the Corona-T-test in discriminating healthy donors from vaccinated and convalescents. (A) Scatter plot shows Corona-T-test result in terms of mean number of spots in duplicates after subtracting the negative control for the Vac_trial (n = 69) and CP_trial (n = 50) cohorts compared to HD-2021 (n = 95). Results in the grey zone, representing samples with 4–6.5 spots (n = 6), were excluded from the ROC-analysis. Mann-Whitney test, p < 0.0001. (B) ROC curve for Vac_trial (AUC = 0.98) and CP_trial (AUC = 0.97) versus HD-2021_trial. (C, D) Number of confirmed HLA-I and -II alleles per individual binding at least one peptide from (C) any protein or (D) S protein (n = 2,210 donors).

We also HLA-genotyped the non-responders (below the grey zone in **Fig. 5A**) from the Vac_trial and CP_trial cohorts to assess possible biases and identify HLA alleles associated with weak response. Among two non-responders in Vac_trial, we observed only 1–2 confirmed HLA alleles presenting epitopes with modest immunogenicity. Two CP_trial non-responders bore three confirmed HLA alleles, which potentially bound 13 and six confirmed peptides, respectively. This is comparable with the median number of recognized epitopes per person in the CP group (**Fig. 4C, D**). Thus, we expect that lower responses, at least in CP individuals, are an intrinsic property rather than a consequence of lacking confirmed HLA alleles. In line with this hypothesis, we observed only modest correlation between the number of confirmed HLA-I alleles and the response to MHC-I peptides in the Vac_trial (r = 0.52, p = 0.0004) cohort, and a number of confirmed HLA-II alleles with MHC-II cross^—^ in the CP_trial (r = 0.4, p = 0.007) cohort. We examined the distribution of the population according to the number of confirmed HLA alleles, and observed that up to 2.8% of Vac_trial—but only 0.1% of CP_trial—lacked any of the confirmed HLA alleles (**Fig. 5C, D**)

### Healthy SARS-CoV-2-exposed individuals demonstrated strong cross-reactivity but no specific response to SARS-CoV-2

During June-July 2020, we recruited an additional cohort of healthy exposed (HE) individuals (n = 37) who were in close contact with COVID-19 patients but did not report any flu-like symptoms and remained IgG- and IgM-negative. The median time of contact was 22 days (Q1 = 10 days, Q3 = 29 days). In comparison to HD-2019, HE individuals demonstrated significantly higher responses to M and S peptides but not to MHC-I and MHC-II cross^—^ peptides (**Fig 6 A, B**). For CP individuals, in contrast, we observed high concordance of positive responses (≥ seven spots) to M, N, or S peptides and MHC-I + II peptides (**Fig. S2A**), with only 2/51 demonstrating a discordant response. Finally, we observed a significant cross-reactive response to the RWY and IED epitopes in the HE cohort that was not seen in the Vac cohort (**Fig. 6C** and **Fig. S3B**). These results show that the HE cohort is substantially different from both CP and HD-2019, suggesting the presence of a cross-reactive T cell response that might have prevented the development of symptomatic disease and T cell priming with other SARS-CoV-2 epitopes from our set.

**Figure 6.**
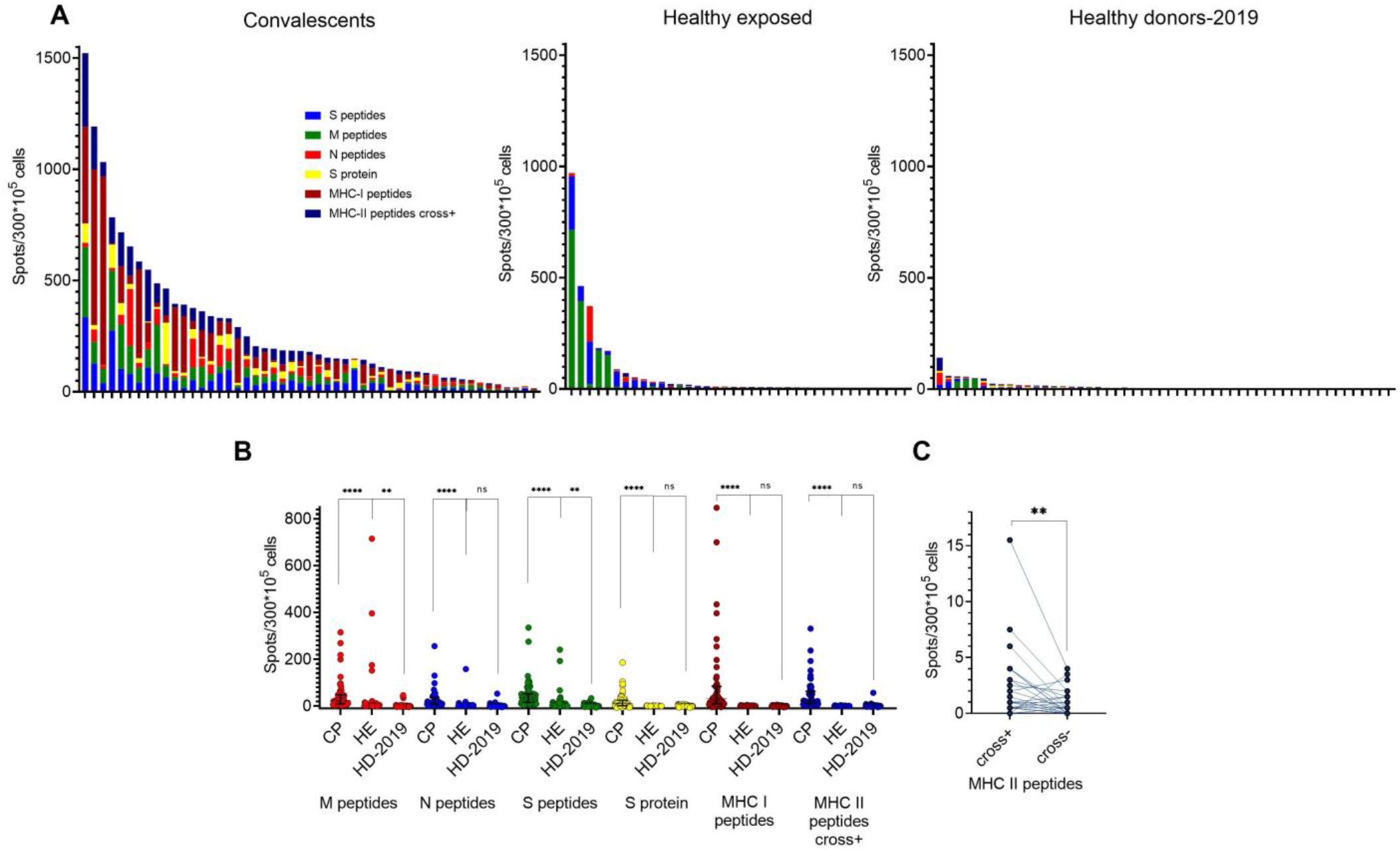
Healthy exposed (HE) are more likely to respond to full-length antigens but not to the MHC-I + II peptides, suggesting rather cross-reactive response than SARS-CoV-2-specific. (A) Response to antigens (mean of two wells after subtracting negative control) in HE (n = 37), CP (n = 51), and HD-2019 (n = 52) cohorts. Each bar represents an individual donor; colors represent particular antigens. (B) Comparison of the response to antigens between cohorts. (C) Difference in responses to MHC-II peptides before and after exclusion of two cross-reactive peptides in HE (Wilcoxon test, p = 0.0045).

## Discussion

Systematic study of the immunogenicity of SARS-CoV-2 T cell epitopes opens the door to vaccine optimization and the utilization of T cell response as an independent measure of immune protection. In the present study, we observed a significant decrease of B*07:02/C*07:02 haplotype frequency in the CP cohort compared to donors from the bone marrow registry, vaccinated and healthy exposed. This HLA bias may be explained by the partial protection of B*07:02 carriers mediated by the T cell response induced by seasonal coronaviruses. At least one epitope presented by this allele is known to be cross-reactive ^41,44^ and immunodominant. Interestingly B*07:02 is a single HLA allele that is characterized by a consistent decrease in the number of predicted epitopes over the course of SARS-CoV-2 evolution ^52^. We have also selected a set of immunogenic and putatively non-cross-reactive SARS-CoV-2 peptides that are predicted to be presented by common HLA-I and -II alleles. Based on analysis of IFNɣ response in 52 pre-pandemic samples (HD-2019), we identified two cross-reactive MHC-II epitopes. Other epitopes (SPR, KLW, and LLY) that have subsequently been proven to be cross-reactive by others ^32,41,44,53^ did not produce a measurable response in our assay, and this could possibly be explained by the low peripheral frequency of specific T cells. In support of that hypothesis, HLAs presenting these cross-reactive epitopes were not enriched in a small subgroup of HD-2021 who were positive based on our Corona-T-test.

We compared the MHC-I + II peptides to commonly-used peptide pools spanning the full-length S, N, and M proteins and to the recombinant S protein. The Vac and CP cohorts did not differ significantly in their response to S, N, and M proteins and to the limited number of peptides in the MHC-I + II set. Surprisingly some vaccinated individuals demonstrated even higher response to MHC-I + II peptides than to S protein-derived peptides, underlining the superior immunogenicity of precise epitopes compared to 15-mer peptides, which require trimming before HLA binding. At the same time, full-length epitopes sporadically induced robust responses in pre-pandemic samples, which makes them ill-suited for measuring SARS-CoV-2-specific T cell response. Carriers of the common HLA allele A*01:01 had a significantly higher response to MHC-I peptides than A*01:01-negative individuals. Moreover, the response to the MHC-I + II set in A*01:01 carriers exceeded the response to the full-length structural proteins, and this can probably be explained to a large extent by the presence of seven confirmed A*01:01 epitopes—including six from non-structural proteins, five of them immunodominant—within the MHC-I + II set. This underlines the notion that in A*01:01 carriers, the majority of the CD8^+^ T cell response might be focused beyond structural proteins, including S protein. Alongside the recent data that the T cell response to early-expressed non-structural proteins may result in protection from infection ^32^, this provides the rationale for designing vaccines that contain such immunodominant peptides. We did not observe an increased response for other HLAs, presenting multiple epitopes (e.g. eight confirmed and three immunodominant epitopes for A*02:01).

T cells from Vac individuals recognized significantly more S protein-derived MHC-I epitopes than those from CP donors, while the total number of recognized MHC-I epitopes from any antigen was, unsurprisingly, higher in the latter group. Alongside the lack of a significant difference between these groups in the magnitude of the response to MHC-I peptides, this allows us to assume higher frequency—and potentially, clonal diversity—of S protein-specific CD8^+^ T cells in vaccinated individuals, resulting from focusing of the response on a single antigen. However, this effect was not replicated in CD4^+^ T cells, and the number of recognized S protein MHC-II epitopes per individual was significantly lower in Vac than in CP. It should be noted that multiple studies have suggested comparable CD8^+^ and CD4^+^ T cell responses at least four weeks after the first dose of vaccine ^9–11,13^. Accordingly, we believe that the negligible PBMC response to recombinant S protein (**Fig. 2A**) and low detection rate of S-protein-specific CD4^+^ T cells after *ex vivo* expansion (**Fig. 4D, S4C**) are associated with late sampling time (median 66 days after the first dose of vaccine). We believe that we did not observe a correlation between sampling time and response to S protein (**Fig. S2D**) because at the earliest sampling time, the peripheral abundance of S-protein-specific CD4^+^ T cells was already below the quantification limit of the ELISpot platform. This may indicate earlier contraction of vaccine-induced S-protein-specific CD4^+^ T cells, or their diminished proliferative potential compared to CD8^+^ T cells.

We used the MHC-I + II peptides identified here as the basis for the ELISpot-based Corona-T-test. This test exhibited 95% accuracy in detecting SARS-CoV-2-specific T cell response in a blinded clinical trial. Intracellular IFNɣ staining performed in parallel was less sensitive, but allowed us to confirm our previous finding that the measured IFNɣ response was largely mediated by CD8^+^ cells. HLA genotyping of non-responders from the Vac_trial and CP_trial cohorts did not show an association between negative response and insufficient coverage of HLA alleles by the epitopes in the set. Instead, this lack of specific T cells was explained by individual variation in the immune response. The rare responders in the HD-2021 group did not have an increased incidence of HLAs presenting the small number of cross-reactive peptides in the MHC-I set, and it is therefore most likely that this group included some seronegative individuals after asymptomatic infection.

HLA restriction was previously reported for most immunogenic MHC-I epitopes (**Table S1**). Nevertheless, due to the large number of immunized donors, we detected some additional—and often less immunodominant—restricting HLAs (*e.g.*, KCY/A*11:01, QUI/A*23:01, and KTF/A*30:01). Several immunogenic epitopes were not, to the best of our knowledge, assigned to their HLA before (*e.g.,* FQP, GRL, SSP, QQQ, FCN, FLL, RYR, VRF, and LQT-derived TYVTQQLI), or at least not with statistical confirmation of their HLA restriction. Five epitopes with two or more detected responses in our study were not reported in previous high throughput screenings ^42,49,54,55^, or were tested only within peptide pools and not individually ^37^. In contrast to MHC-I epitopes, MHC-II epitopes were mostly promiscuous in their HLA-binding (**Fig. 4B**). Most of the CD4^+^ response was focused on non-S-protein epitopes. Compared to the most comprehensive screening performed to date date ^49^, we identified an additional 11 MHC-II epitopes, and statistically confirmed HLA associations for nine of them. Five precise epitopes were predicted from immunogenic peptides ^50,56^; six others were annotated in IEDB epitopes of SARS-CoV-1. We also confirmed two MHC-I epitopes within MHC-II epitopes.

Although there are several kits aimed at detecting SARS-CoV-2-specific T cells, we believe that the strategy of epitope selection is critical for discriminating pre-existing cross-reactive immunity from specific immunity. Since we confirmed that the MHC-I + II peptide set can accurately discriminate SARS-CoV-2-induced and cross-reactive immunity, we next characterized the HE cohort of individuals without clinical or laboratory signs of COVID-19 after prolonged contact with COVID-19 patients. Surprisingly, we saw virtually no response (1 of 37) to MHC-I + II peptides, versus relatively frequent and robust responses to S, N, and M peptides, as well as significant responses to the cross-reactive IED and RWY peptides. The discordant responses in this group contrasted sharply with the CP cohort, in which responses to full-length antigens correlated strongly with response to the MHC-II set (**Fig. S2B**). This is best explained by the protective effect of pre-existing cross-reactive T cell immunity in this cohort. Notably, we initially recruited two patients with a low-level IgM anti-S protein response, who were subsequently excluded using a more sensitive commercial kit. These donors, however, demonstrated an easily detectable MHC-I + II response (18 and 54 spots), suggesting that the reported peptide set could even be used to diagnose asymptomatic CP with a negligible humoral response. Based on these results, we believe that the described set of SARS-CoV-2 epitopes offers a sensitive and specific tool for the detection of COVID-19 or vaccination-induced (but not cross-reactive) T cell response.

## Materials and Methods

### Donors

*The following describes donors for the non-clinical-trial component of this study.* Healthy donors pre-pandemic (HD-2019, n = 52) included 29 PBMC samples containing at least 20 х 10^6^ cells cryopreserved before December 2019 and 23 cryobags obtained via mononuclear cell apheresis. The CP samples (n = 51) included PBMCs from patients who had COVID-19 in February–May 2020 (n = 43) or January–June 2021 (n = 8), which were collected and frozen within 20–70 days after the onset of symptoms. Assessments of individual epitope immunogenicity were performed on 48 of these samples. Vac PBMCs (n = 45) were collected and frozen 23–65 days after receiving the second dose of the Sputnik-V (Gam-COVID-Vac) vaccine from donors who had a negative antibody tests no later than four weeks before the first shot and no self-reported COVID-like symptoms. ELISpot measurements were performed on 43 of these samples. The HE cohort (n = 37) of samples were from people who had close contact with a patient with active COVID-19 (same household or “red zone” medical workers with multiple negative RT-PCR tests) but who were themselves without COVID-19 symptoms and without detectable IgG and IgM anti-S protein antibodies.

*The clinical trial of the Corona-T-test kit* was conducted at Dmitry Rogachev National Medical Research Center of Pediatric Hematology, Oncology, and Immunology in Moscow, Russia. Donors were recruited by a separate center, DNKOM Laboratory, where the samples were collected and blinded. Blood sampling took place between July and September 2021. There were 220 participants in three cohorts: 1) 101 healthy (HD-2021) patients, 2) 50 recent convalescents after COVID-19 (CP_trial), and 3) 69 vaccinated with Sputnik V (Vac_trial). All samples were pretested for antibodies against SARS-CoV-2, and inclusion criteria and antibody tests are shown in the table below.

| Cohort | Inclusion criteria | Serology testing |
| --- | --- | --- |
| Healthy patients (HD-2021; n = 101) | No self-reported COVID-19 infection | 1. IgM to SARS-CoV-2 detection: “SARS-CoV-2-IgM-ELISA-BEST”, AO “Vector-Best”, r.p. Koltsovo, Russia. Ref #D-5502, Registration certificate № RZN 2020/10389<br>2. IgG to SARS-CoV-2 S protein |
|  | No vaccination against COVID-19 |  |
|  | No IgM or IgG antibodies against SARS-CoV-2 by three tests | detection: “SARS-CoV-2-IgG-quantity-ELISA-BEST”, AO “Vector-Best”, r.p. Koltsovo, Russia. Ref #D-5505, Registration certificate № RZN 2021/14458<br>3. IgG to SARS-CoV-2 N protein detection: “SARS-CoV-2 IgG Reagent Kit”, ARCHITECT, Abbott Laboratories, EU. Registration certificate № RZN 2020/11359. |
| Recent COVID-19 convalescents (CP_trial; n = 50) | Self-reported COVID-19 infection | IgG to RBD domain of SARS-CoV-2 S protein detection: “SARS-CoV-2-IgG-ELISA”, “National Medical Research Center for Hematology”, Moscow, Russia. Ref #K153G, Registration certificate № RZN 2020/10815 |
|  | PCR confirmed COVID-19 in a period 14–45 days before recruitment |  |
|  | Detectable IgG antibodies against SARS-CoV-2 |  |
| Vaccinated with “Sputnik V” (Vac_trial; n = 69) | Documented confirmation of vaccination against SARS-CoV-2 in a period 7–14 days after second vaccination before recruitment | IgG to RBD domain of SARS-CoV-2 S protein detection: “SARS-CoV-2-IgG-ELISA”, “National Medical Research Center for Hematology”, Moscow, Russia. Ref #K153G, Registration certificate № RZN 2020/10815 |
|  | Detectable IgG antibodies against SARS-CoV-2 |  |

Written informed consent was obtained from all patients. All patients were insured during the clinical trial. Blood sampling and PBMC separation were performed with Vacutainer CPT Cell Preparation Tubes with sodium heparin^N^ for the separation of mononuclear cells from whole blood (BD, Belgium). Each PBMC sample was divided into two parts and used for measuring T cell response with the Corona-T-test kit and by flow cytometry with intracellular IFNɣ staining.

### PBMC isolation and HLA genotyping

30 mL of venous blood from healthy donors, vaccinated individuals, and recovered COVID-19 patients was collected into EDTA tubes (Sarstedt, Germany) and subjected to Ficoll (Paneco, Russia) density gradient centrifugation (400 x *g*, 30 min). Isolated PBMCs were washed with PBS containing 2 mM EDTA twice, counted, cryopreserved in 7% DMSO and 93% heat-inactivated fetal bovine serum (Capricorn Scientific, Germany), and stored in liquid nitrogen until used in the assays.

### HLA genotyping

For the Vac cohort, HLA genotyping was performed with the HLA-Expert kit (DNA-Technology LLC, Russia) through the amplification of exons 2 and 3 of the HLA-A/B/C genes and exon 2 of the HLA-DRB1/3/4/5/DQB1/DPB1 genes. Prepared libraries were run on an Illumina MiSeq sequencer using a standard flow-cell with 2 x 250 paired-end sequencing. Reads were analyzed using HLA-Expert Software (DNA-Technology LLC, Russia) and the IPD-IMGT/HLA database 3.41.0 (10.1093/nar/gkz950).

For CP_trial, Vac_trial, HD-2021, and CP donors, HLA-genotyping was performed using the One Lambda ALLType kit (Thermo Fisher Scientific, USA), which uses multiplex PCR to amplify full HLA-A/B/C gene sequences, and from exon 2 to the 30 UTR of the HLA-DRB1/3/4/5/DQB1 genes. Prepared libraries were run on an Illumina MiSeq sequencer using a standard flow-cell with 2 x 150 paired-end sequencing. Reads were analyzed using One Lambda HLA TypeStream Visual Software (TSV), version 2.0.0.27232, and the IPD-IMGT/HLA database 3.39.0.0. Two donors (p1305, p1329) were HLA genotyped by Sanger sequencing for loci HLA-A, B, C, DRB1 and DQB1, using Protrans S4 and Protrans S3 reagents, respectively. The PCR products were prepared for sequencing with BigDye Terminator v1.1. Capillary electrophoresis was performed on a Genetic Analyzer Nanophore05.

### IFNɣ ELISpot (for the non-clinical trial part of this work)

Cells from cryotubes or blood cryobags were thawed, rested 4–24 hours in CTL-test media (CTL, USA), counted, and plated in human IFNɣ ELISpot plates (CTL, USA) at 3 х 10^5^ PBMCs per well and incubated with different sets of antigens: 1 μM peptides MHC-I, 1 μM MHC-II cross^+^, 1 μM MHC-II cross^—^, 10 μg/mL full-length S protein, or 1 μg/mL commercial peptide pools covering S, N, and M proteins (Miltenyi Biotech, Germany). S protein-derived peptides including a combination of PepTivator SARS-CoV-2 Prot_S, PepTivator SARS-CoV-2 Prot_S1, and PepTivator SARS-CoV-2 Prot_S+. For the positive control we used 40 ng/mL PMA (P8139-1MG, Sigma; USA) and 400 ng/mL calcium ionophore (C7522-1MG, Sigma; USA), with 0.02% isopropanol plus 0.02% DMSO as a negative control. After 18–20 hours of incubation at 37 °C, 5% CO_2_, the plates were developed according to the manufacturer’s guidelines. Spots were counted using the CTL ImmunoSpot Analyzer (CTL, USA). T cell responses were considered positive when more than seven spots (mean of duplicates) were detected after subtracting the negative control. Samples with >17 spots in the negative control or < 50 spots in the positive control were excluded from the analysis.

### T cell expansion

PBMCs from cryotubes or blood cryobags were thawed, rested 4–24 hours in CTL-test medium, counted, and used for rapid *in vitro* expansion. 2–4 x 10^6^ cells per well were plated in 24-well plates in RPMI 1640 culture medium supplemented with 10% normal human A/B serum, 1 mM sodium pyruvate, 2 mM GlutaMAX Supplement (Thermo Fisher Scientific, USA), 25 ng/mL IL-7, 40 ng/mL IL-15, and 50 ng/mL IL-2 (Miltenyi Biotec, Germany) at a volume of 1 mL/well. The initial full set of peptides (final concentration of each = 10 μM) were added on day 0. On day 3, 1 mL of supplemented medium without peptides was added to each well (final volume 2 mL). Half of the medium was replaced on days 5, 7, 10. On days 10 and 11, an aliquot of cell suspension was used for anti-IFNɣ ELISA with pooled peptides and individual peptides respectively. On day 13, cells were sampled for flow cytometry analysis. In order to maintain detectable IFNɣ secretion, a quarter of the medium was replaced with supplemented medium on days 11 and 13.

### Cell stimulation with peptide pools

After 10 days of expansion, an aliquot of cell culture was washed twice in 1.5 mL of PBS, then transferred to AIM-V medium (Thermo Fisher Scientific, USA), plated at 10^5^ cells per well in 96-well plates, and incubated overnight (12–16 hours) with the peptide pools. 0.04% DMSO and 0.04% isopropanol was used as a negative control, with 40 ng/mL PMA and 400 ng/mL calcium ionophore as positive control. On day 11, the culture medium was collected and tested for IFNɣ as described below in the “Anti-IFNɣ ELISA” section. On days 11–12, we stimulated the cells as described above individually with each peptide (2 μM) from the pools. Only peptides predicted to bind to each individual’s HLA were tested. Finally, on day 13, we stimulated the MHC-II peptide-expanded cells with the MHC-II peptides that generated a positive response with ELISA for flow cytometry experiments.

### Anti-IFNɣ ELISA

96-well high-protein-binding ELISA plates (Shanghai Meikang Biological Project, KH-M-02, China) were coated with 100 μL per well of 0.01 mg/mL anti-IFNɣ antibody (Hytest, clone GF1) in 100 mM bicarbonate/carbonate (pH 9.6). After 14 h, the plates were washed once with 250 mL PBS + 0.1% Tween 20 (PBST) and blocked with 100 mL of 1% hydrolyzed casein (XEMA, Russia) in PBS for 3 h at room temperature, dried, and stored sealed at 4 °C.

Culture plates were centrifuged for 3 min at 700 x *g*, and 100 µL of the medium was transferred to the ELISA plates. Plates were incubated for 1.5 h at 37°C on a rocking platform. The plates were washed thrice with PBST, and then 100 µL of 0.1 µg/mL biotinylated anti-IFNɣ antibody (R&D Systems, USA) was added and incubated for one hour at 37°C on a rocking platform. The plates were washed thrice with PBST, and then 100 µL of Streptavidin-HRP (XEMA, Russia) was added and incubated for 0.5 h at 37°C on a rocking platform. Finally, the plates were washed five times with PBST, and 100 µL of 3,3’,5,5’-tetramethylbenzidine substrate (XEMA, Russia) was added to each well. 15 min later, 100 µL of 1 M H_2_SO_4_ was added as a stop solution, and the optical density (OD) was measured at 450 nm on a MultiScan FC (Thermo Fisher Scientific, USA) instrument. Each plate included standards corresponding to 0, 15, 120, and 7,700 pg/mL of IFNɣ to control the sensitivity and linearity.

Test wells with peptides where the ratio OD_450_test_well_/OD_450_negative control_ ≥ 1.25 and the difference OD_450_test_well_ – OD_450_negative control_ ≥ 0.08 were considered positive. Peptides with a ratio between 1.25 and 1.5 were tested again up to three times as biological replicates to ensure the accuracy of their response. Peptides with two or three positive results were considered positive. If such peptide was not repeatedly tested, it was considered negative. If ELISA results conflicted with flow cytometry data (for MHC-II peptides), the peptide was considered non-immunogenic.

### Flow cytometry

After 13 days of expansion, an aliquot of cell suspension was washed twice in 1.5 mL of PBS, then resuspended in AIM-V medium with 1.0 µg/mL brefeldin A (GolgiPlug, BD Biosciences; USA). Cells were plated at 10^5^ per well in a 96-well polypropylene V-bottom plate and incubated at 37 °C for ∼5 h with MHC-II peptides that were identified as positive in the previous ELISA assay. 0.04% DMSO and 0.04% isopropanol were used as a negative control, and 40 ng/mL PMA and 400 ng/mL calcium ionophore were used as positive control. Stimulation was stopped by washing with PBS plus 0.5% BSA (Sigma-Aldrich, USA) and 2 mM EDTA. Surface staining was performed for 10 min with Alexa Fluor 750 NHS Ester (Thermo Fisher Scientific, USA) in 100 μL PBS at room temperature, followed by two washes with 10% FBS diluted with PBS and staining with anti-CD3-FITC (SK7; cat.345764 Sony, USA), anti-CD4-PE (SK3; cat.345769; BD Biosciences, USA), and anti-CD8-APC (SK-1; cat.345775; BD Biosciences) for 20 min at 4 °C. Fixation and permeabilization were performed with BD Cytofix/Cytoperm fixation and permeabilization solution (BD Biosciences, USA), according to the manufacturer’s protocol followed by IFNɣ-BV421 (B27; cat.562988; BD Biosciences, USA) staining for 20 min at 4 °C. All samples were analyzed with a MACSQuant Analyzer 10 (Miltenyi Biotec, Germany). Instrument performance was monitored prior to every measurement with MACSQuant Calibration Beads (Miltenyi Biotec, Germany). The acquired data was processed by FlowJo software (version 10.6.2., Tree Star, Ashland, OR). The percentage of IFNɣ-positive cells was calculated in the CD3^+^CD8^+^ and CD3^+^CD4^+^ gate. The difference or ratio (fold-change) of the percentage of IFNɣ-positive cells incubated with peptide and with negative control was calculated. Peptides with a ratio >2 were considered positive for CD4^+^ recognition, or with a ratio >3 for CD8^+^ recognition due to a higher background % of CD8^+^IFNɣ^+^ cells. If the total number of CD8^+^ cells was lower than 5000 cells, this well was not analyzed for the CD8^+^ response. In case of conflicting results in ELISA and flow cytometry the peptide was considered negative.

### Anti-SARS-CoV-2 ELISA

ELISA kits for the detection of anti-RBD IgG (K153, National Research Center for Hematology, Russia) and SARS-CoV-2-IgМ-EIA-BEST (D-5502, Vector Best, Russia) for the detection of IgM antibodies to full-length S protein were used according to the manufacturers’ instructions.

### Assessing epitopes homology

To assemble the control peptide pool, IEDB was queried for epitopes with positive MHC binding and a minimum of two positive T cell assays using ‘Severe acute respiratory syndrome-related coronavirus’ as ‘Organism’ on 11 October 2021. MHC-I and MHC-II peptides used in this study were excluded from the IEDB epitope pool. Alignments were performed using best global-local alignment by the ‘pairwiseAlignment’ (bioPython) function for four ORFs shared by all five strains: orf1ab, S, N, and M, allowing amino acid substitutions with similar biochemical properties (1, 2) and low penalties for gap opening and extension (−0.5). The segments of the alignments corresponding to the given SARS-CoV-2 peptide were further aligned again with high gap penalties (−10/-1) followed by calculation of the similarity score ^57,58^nand identity score.

### HLA and epitope selection

We selected the HLA list based on the most-presented HLA among the CP cohort:

**HLA-I:** A*01:01; A*02:01; A*03:01; A*11:01; A*23:01; A*24:02; A*25:01; A*30:01; A*32:01; B*07:02; B*08:01; B*13:02; B*15:01; B*18:01; B*27:05; B*35:01; B*38:01; B*40:01; B*44:02; B*44:03; B*51:01; C*01:02; C*03:04; C*04:01; C*05:01; C*06:02; C*07:01; C*07:02; C*12:03; C*15:02

**MHC-I peptides:** The core set comprised two epitopes from Shomuradova *et al.* and 25 minimally/non-cross-reactive epitopes from the publications listed in **Fig 1A**. 67 epitopes from Snyder *et al.* were chosen with the aim of selecting individual epitopes instead of peptides within a peptide pool and balancing predicted HLA coverage, minimal cross-reactivity (based on the number of non-naive expansions from healthy donors assigned to a specific epitope/peptide group), and higher immunogenicity (based on the number of CP with detected TCR sequences assigned to a specific epitope/peptide group). For the core set and all of the peptides from Snyder *et al.*, we predicted HLA binding, using NetMHCPan4.1 with standard thresholds, strong and weak binders were considered as potential epitopes for a given HLA allele.

**HLA-II**: DRB1*01:01; DRB1*07:01; DRB1*11:01; DRB1*11:04; DRB1*13:01; DRB3*01:01; DRB3*02:02; DRB4*01:01; DQB1*02:01; DQB1*03:01; DQB1*03:02; DQB1*05:01; DQB1*06:03

**MHC-II peptides**: These were minimally cross-reactive and immunogenic peptides from the publications listed in **Fig. 1A**. Thereafter, we searched for the exact MHC-II epitopes from those peptides and predicted their binding to the selected HLA-II epitopes assigned to SARS-CoV-1 in IEDB and four peptides with predicted high binding promiscuity (> 7 HLA-II alleles). HLA binding was predicted using both NetMHCIIpan 3.2 ^59^ and Neon MHC2 ^60^ for MHC-II peptides using standard thresholds. Strong binders, weak binders, or peptides with discordant NetMHCIIpan and Neon MHC2 predictions were considered as potential binders and were tested in donors bearing those HLAs.

### Peptides and mixes

Peptides (at least 95% purity) were synthesized either by Peptide 2.0 Inc. (VA, USA) or by the Laboratory of Ligand-Receptor Interactions (Zhmak, M. N.) from the Shemyakin-Ovchinnikov Institute of Bioorganic Chemistry RAS (Moscow, Russia). Peptides containing Cys and/or Met were diluted in 50% isopropanol in PBS at concentrations of up to 10–25 mM. Other peptides were diluted in DMSO (Sigma) at up to 30–40 mM. When peptide pools were prepared, peptides containing Cys/Met were not mixed with peptides in DMSO. All individual peptides and mixes were aliquoted for single-use and stored at −80°C.

For identification of cross-reactive peptides, matrix pools were prepared (20 matrices for MHC-I and 11 for MHC-II), with each peptide present only in two pools. IFNɣ production in wells containing both pools indicated the specificity of the corresponding peptide (**Fig. S1F**). We used the following mixes in our work (each presented by two submixes, diluted in DMSO or isopropanol):

1. For the ELISpot assay (final concentration in medium = 1 μM): i) MHC-I peptides, containing all MHC-I peptides, ii) MHC-II peptides (cross+) – all MHC-II peptides, iii) MHC-II peptides cross^—^ – MHC-II peptides without RWY and IED, iv) MHC-I + II _IVD – pre-mixed, aliquoted and lyophilized set of 115 peptides without RWY, IED, and SEL (which was excluded due to its physicochemical properties hindering synthesis), and v) matrix mixes (20 with MHC-I peptides and 11 with MHC-II peptides)
2. For T cell expansion (final concentration in medium = 10 μM): the full set of peptides was split into five standard mixes (Expansion pools, **Table S1**).
3. For the first step of epitope identification in anti-IFNɣ ELISA 5*(∼5) = 26 standard mixes (ELISA-pools, **Table S1**) corresponding to the composition of the five standard mixes for T cell expansion.

### Quantification and statistical analysis

All data comparisons were performed using GraphPad Prism 8, Python 3.2, and FlowJo 10 software. Statistical analyses were performed using Spearman correlation and Mann-Whitney tests for unpaired comparisons or Kruskal-Wallis test followed with Dunn’s post hoc test for comparing multiple groups. For paired measurements, we used the Wilcoxon test. For detection of the statistical association between HLA alleles and MHC-II epitopes or non-S protein-derived MHC-I epitopes, we used only the CP cohort; for HLA-I alleles and S protein-derived MHC-I epitopes, we used both CP and Vac cohorts. The number of responders among HLA carriers and the number of responders among individuals without this HLA were compared with Fisher exact test, with p-values < 0.05 considered significant.

### Measuring SARS-CoV-2 specific T cell response with Corona-T-test (for the clinical trial)

The detection of SARS-CoV-2-specific T cells was performed by Corona-T-test – a single-color enzymatic ELISpot kit for IFNɣ detection produced by the National Research Center for Hematology. After washing cells with sterile RPMI-1640 media twice, cells were counted in a hemocytometer (Sysmex XS-1000i, Sysmex Corporation, Japan) and resuspended in serum-free AIM-V + AlbuMAX BSA (Thermo Fisher Scientific, USA) medium to a concentration of 6 x 10^6^ /mL, and then 3 x 10^5^ cells were loaded per well. The SARS-CoV-2 antigens (MHC-I + II _IVD) were used at a final concentration of 1 μM/mL. Phytohemagglutinin (PHA) was used as a positive control at a concentration of 10 μg/mL. The final volume of each well was 150 μL. All manipulations with cells and antigen dilutions were performed in serum-free media in sterile conditions. We used four wells for each PBMC sample: negative control stimulated with AIM-V medium, SARS-CoV-2-antigen-induced stimulation in duplicate, and positive control with PHA stimulation. Plates were incubated 16–18 hours at 37 °C, 5% CO_2_. The next day, the assay was developed according to the manufacturer’s guidelines, with spots counted using the automated ImmunoSpot Series 5 UV Analyzer (CTL, USA) using automated software. Results were considered valid if the number of spots was < 10/well in negative controls and > 100/well in positive controls. Non-specific activation in negative controls was subtracted from the average of the two stimulated sample wells. Responses with < 4 spots were considered negative and > 6.5 spots were considered positive. Grey zone samples with 4–6.5 spots were considered inconclusive, requiring repeated testing, and such samples (n = 6) were excluded from the accuracy analysis.

### Flow cytometry with the intracellular IFNɣ staining protocol

For flow cytometry assessment, PBMCs were separated as described previously and rested overnight at 7.5 x 10^6^ cells/mL in AIM-V + AlbuMAX (BSA) medium at 37°C with 5% CO_2_. The next day, 200 μL of cell suspension was added to each well of a 96-well polypropylene V-bottom plate. All samples received 0.25 μL of brefeldin A at the beginning of incubation. For the immune stimulation, 1 μM MHC-I + II_IVD peptides was added. 1.5 μg/mL PHA (Capricorn Scientific GmbH, Germany) was used as a positive control. Non-stimulated cultures were used to assess spontaneous intracellular levels of cytokines. The cells were incubated for 4 hours at 37 °C, 5% CO_2_ and then transferred into 12 x 75 mm plastic tubes at 1.5 x 10^6^ cells per tube and washed with 2 mL MACS PBS/EDTA Buffer without Ca^2+^ or Mg^2+^ (Miltenyi Biotec, Germany) with 0.5% heat-inactivated MACS BSA (Miltenyi Biotec, Germany), PBS/BSA/EDTA. The cells were centrifuged at 350 x *g* for 5 min and then stained. Cell surface staining of T cells was done in 0.1 mL PBS/BSA/EDTA for 15 min with FITC-conjugated anti-CD3 (clone SK7, BD Biosciences, USA), PE-Cy7 anti-CD8 (clone SK1, BD Biosciences, USA), VioGreen anti-CD4 (clone REA623, Miltenyi Biotech, Germany), and 7-amino-actinomycin D (7-AAD, Miltenyi Biotech, Germany) in the dark at room temperature. Fixation and permeabilization were performed with BD Cytofix/Cytoperm according to the manufacturer’s protocol, and intracellular staining was carried out for 30 min in the dark at room temperature with APC anti-IFNɣ (clone B27, BD Biosciences, USA). Cells were analyzed on a CytoFLEX (Beckman Coulter) flow cytometer. Instrument performance was monitored daily with CytoFLEX Daily QC Fluorospheres (Beckman Coulter, USA). Individual populations were gated according to forward scatter, side scatter, and specific markers, and the data were subsequently analyzed with CytExpert software (Beckman Coulter, USA). Dot plots representing anti-CD3 versus anti-IFNɣ were carried out to establish CD3^+^IFNɣ^bright^ lymphocyte gates. Identical dot plots were generated for CD8^+^IFNɣ^bright^ and CD4^+^IFNɣ^bright^ cells. Typically, 300,000 events were acquired in the gating CD3^+^ region. Non-specific activation in unstimulated controls was subtracted from stimulated samples to account for specific activation.

## Data Availability

All data produced in the present study are available upon reasonable request to the authors

## Notes

### Competing Interest Statement

The authors have declared no competing interest.

### Funding Statement

The work was supported by a Russian Science Foundation grant 20-15-00395 (G.A.E.)

### Author Declarations

Ethics committee of the National research center for Hematology gave ethical approval for this work

