## Supplementary material for "Immunogenic epitope panel for accurate detection of non-cross-reactive T cell response to SARS-CoV-2": Figures S1-S6

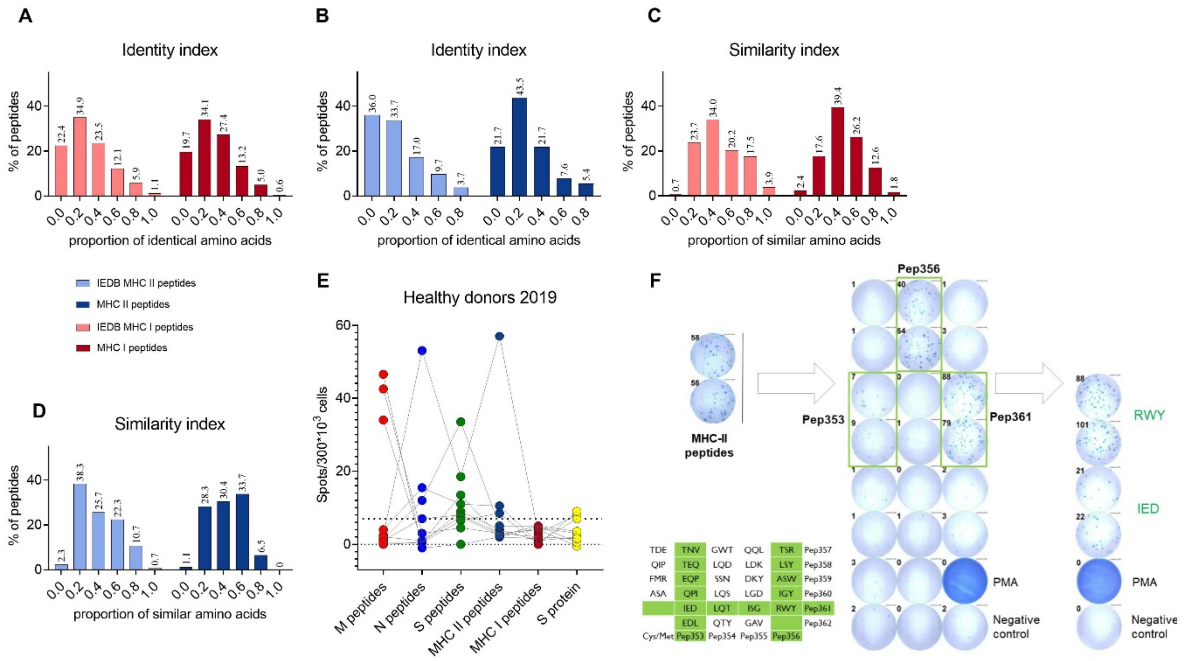


**Figure S1**. (A–D). Identity (A and B) and similarity (C and D) of the MHC-I (A and C) and MHC-II (B and D) peptides, aligned to common cold coronavirus (229E, NL63, HKU1, OC43) proteins. Peptides from IEDB were used as a control. (E) Response to the indicated antigens among healthy donors (grouped for each donor). Responders to at least one antigen are shown. Dots show the mean of two duplicates with negative control subtracted. Threshold for positive result (7) is indicated by the dotted line. (F) The strategy for identifying cross-reactive MHC-II epitopes. This particular donor responded to MHC-II peptides. Identification was performed using matrix pulls; the peptide composition of those pools is indicated in the table at lower left. Green marks matrix pools that evoked a response. When we tested the same donors' peripheral blood mononuclear cells (PBMCs) against the individually-identified peptides (right) we observed a matching response.

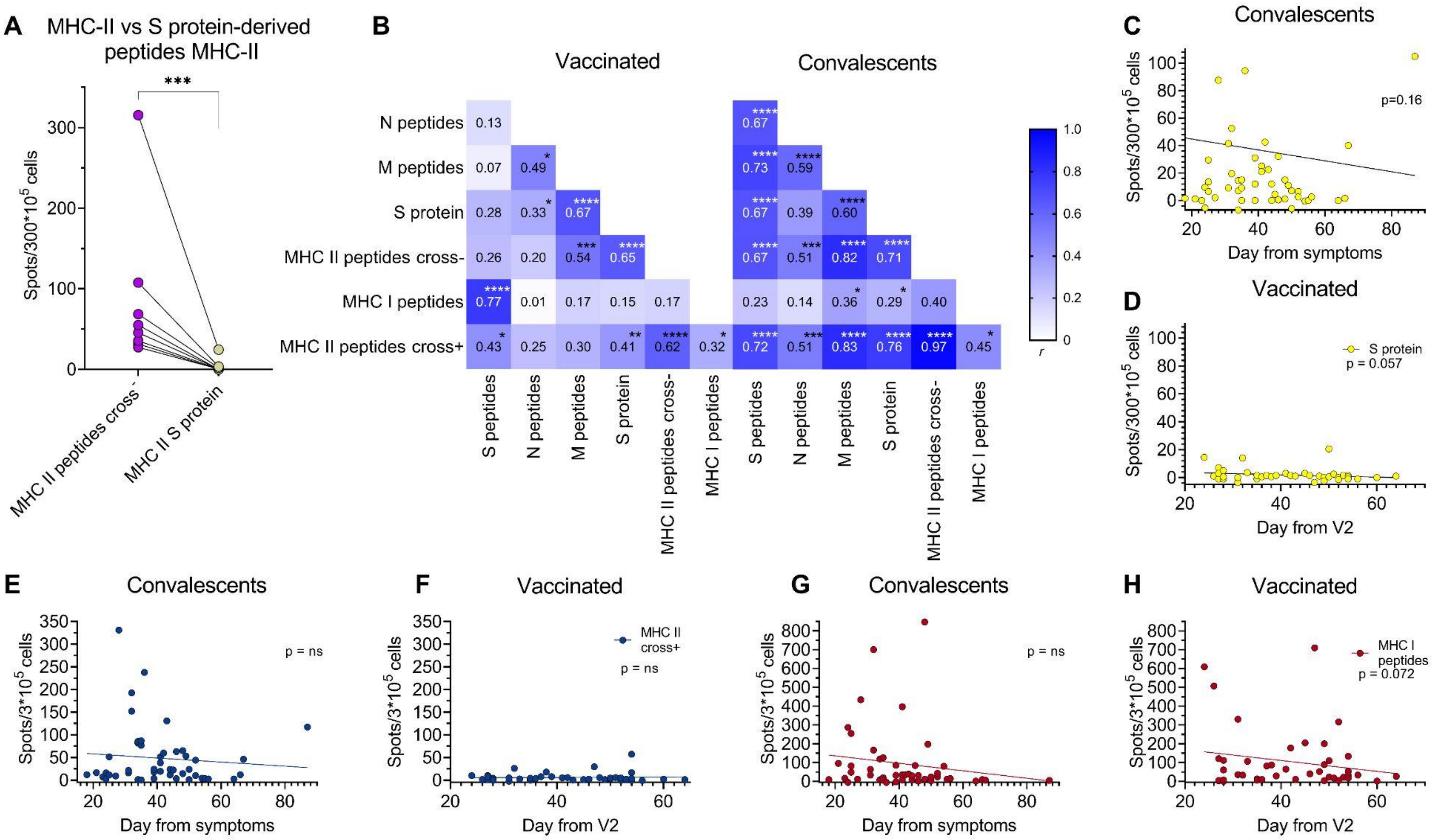


**Figure S2.** (A) Difference in response to MHC-II peptides versus S protein-derived MHC-II peptides (Mann-Whitney test, p=0.002). (B) Correlation between responses to different antigens in Vac and CP (Pearson correlation, *p < 0.05, **p < 0.01, ***p < 0.001, ****p < 0.0001). (C–H) Influence on time elapsed from symptoms or boost vaccination (V2) on the magnitude of response to recombinant S protein in CP (C) and Vac (D), MHC-II peptides in CP (E) and Vac (F), and MHC-I peptides in CP (G) and Vac (H) (linear regression).

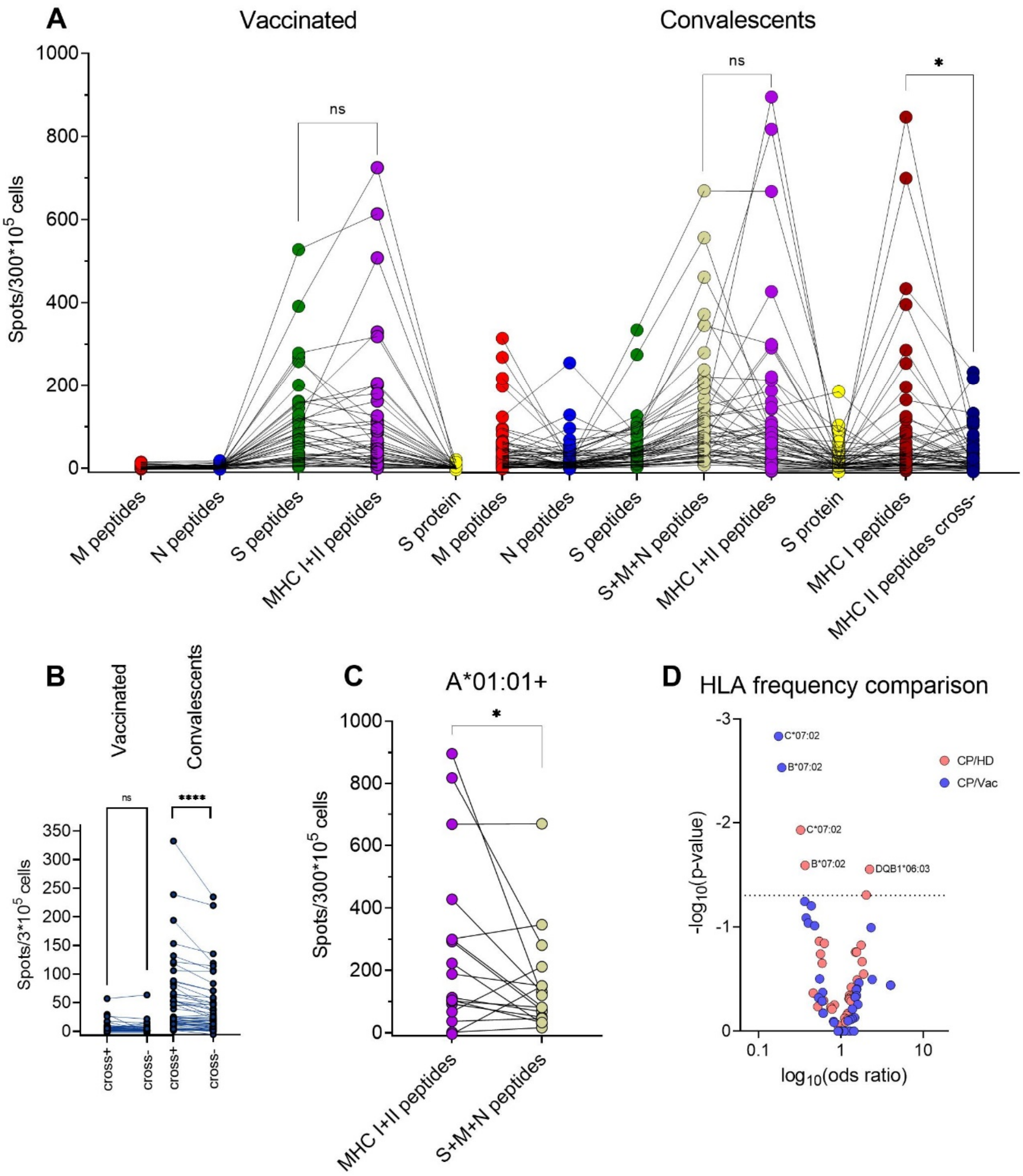


**Figure S3.** (A) Response to antigens and peptide sets grouped by individuals. (B) Difference in responses to MHC-II peptides before and after exclusion of two cross-reactive peptides (Wilcoxon test, $p < 0.0001$) (C) Response to antigens among A*01:01$^{+}$ convalescents (Wilcoxon test, $p = 0.046$). (D) Volcano plot shows the difference between HLA allele frequencies in groups. The x-axis denotes the decimal logarithm of the odds ratio of the given allele carriers and non-carriers between different cohorts (HD - bone marrow donors, n=2210). Y-axis denotes the negative decimal logarithm of the p-value. P-value = 0.05 is depicted by the dotted line (Fisher exact test, statistically significant values are annotated).

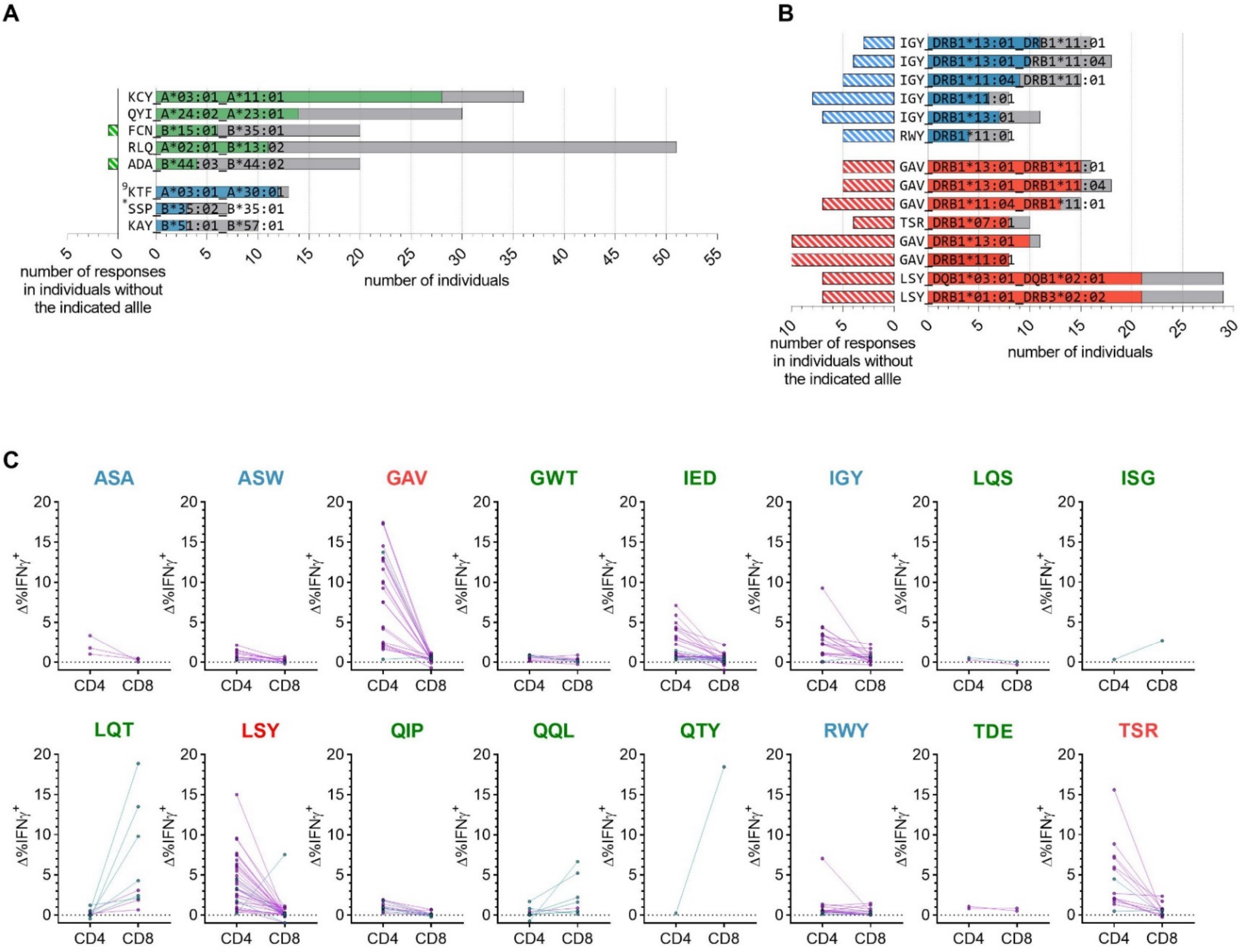


**Figure S4.** (A, B) Number of responses to each peptide in individuals with (right) or without (left) the indicated (A) HLA-I or (B) -II alleles. Data are presented as in **Figure 4A, B**. (C) Flow cytometric analysis of the phenotype of T cells responding to MHC-II peptides for each peptide (antigens labeled green for S, red for M, and blue for N) in Vac (blue) and CP donors (rose). Plots show differences in the % of CD4 or CD8 IFNɣ+ T cells between peptide-stimulated cells and negative controls.

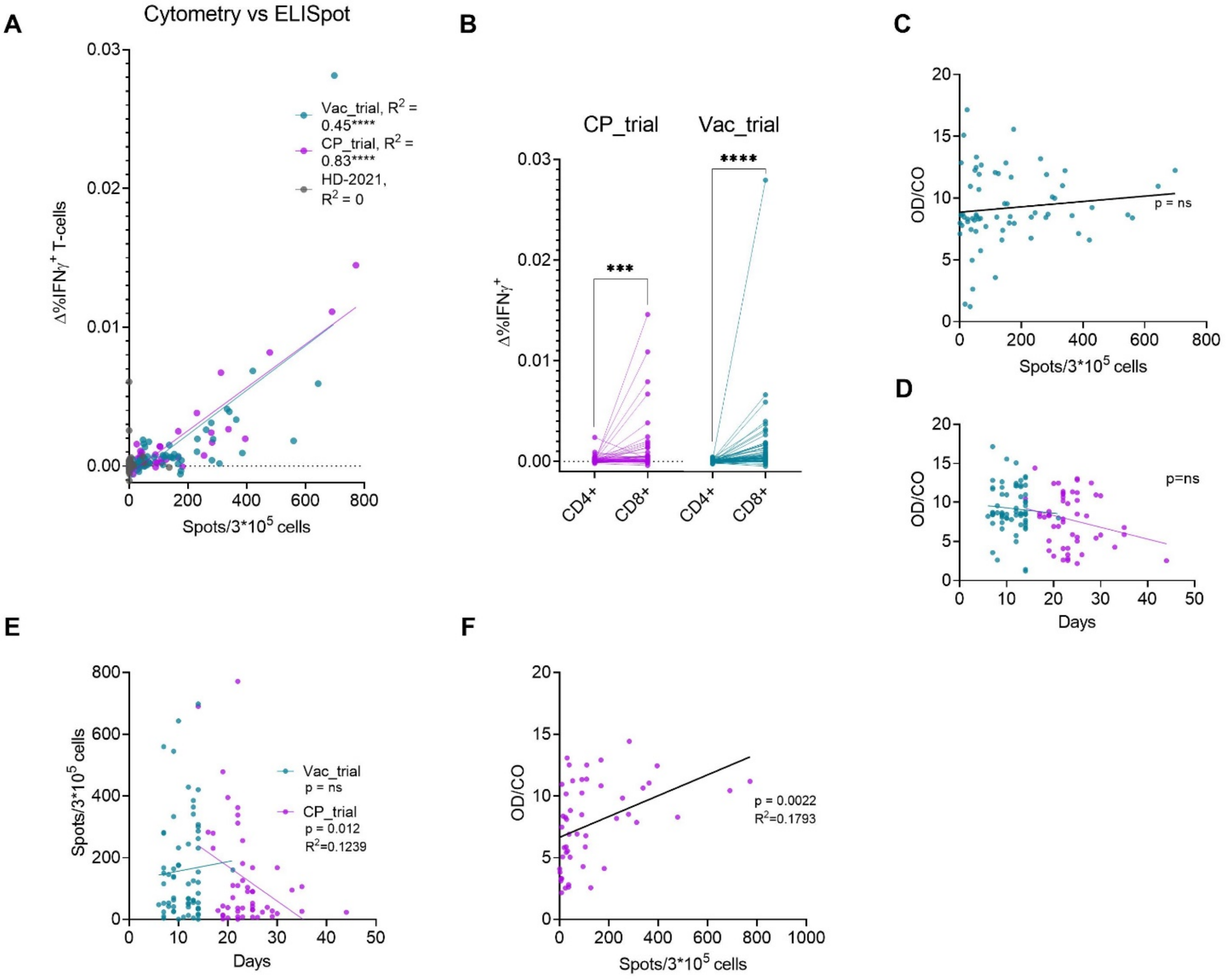


**Figure S5**. (A) Scatter plot shows response of Vac_trial (n = 60), CP_trial (n = 48), and HD-2021 (n = 88) cohorts as measured by Corona-T-test (number of spots; x-axis) versus flow cytometry (sum of %CD4$^+$ IFNɣ$^+$ and %CD8$^+$ IFNɣ$^+$ after subtracting background; y-axis). Linear regression, F-statistic, ****p < 0.0001. (B) Impact of CD4$^+$ and CD8$^+$ on the total response. Plot shows the difference between the peptide-stimulated wells and negative control in IFNɣ$^+$ cells gated as CD4$^+$ or CD8$^+$ cells. Wilcoxon test, ***p = 0.0009, ****p < 0.0001. (C) Correlation between Corona-T-test response and time since V2 (boost vaccination) in Vac_trial individuals. (D, E) Correlation between time since V2 in Vac_trial or disease onset in CP-trial and (D) humoral or (E) T cell response. OD/CO represent the ratio of $OD_{450}$ of the sample to the test cut-off. (F) Correlation between Corona-T-test response and time since disease onset in convalescents.

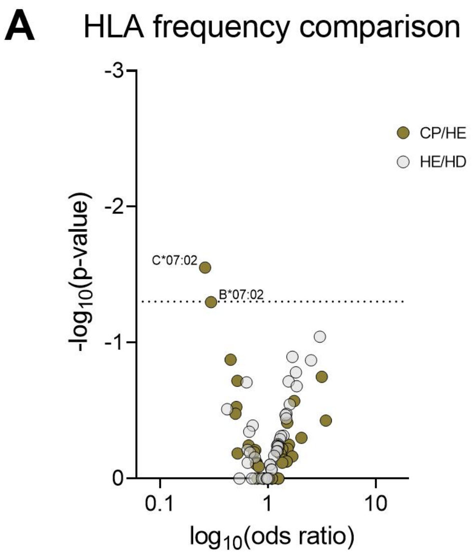


**Figure S6**. (A) Volcano plot shows the difference between HLA allele frequencies in groups. The x-axis denotes the decimal logarithm of the odds ratio of the given allele carriers and non-carriers between different cohorts (HE – healthy exposed, CP – convalescents, HD - bone marrow donors, n=2210). Y-axis denotes the negative decimal logarithm of the p-value. P-value = 0.05 is depicted by the dotted line (Fisher exact test, statistically significant values are annotated).
